# Development of Accurate Long-lead COVID-19 Forecast

**DOI:** 10.1101/2022.11.14.22282323

**Authors:** Wan Yang, Jeffrey Shaman

**Affiliations:** Department of Epidemiology, Mailman School of Public Health, Columbia University, New York, NY, USA; Herbert Irving Comprehensive Cancer Center, Columbia University Medical Center, New York, NY, USA; Department of Environmental Health Sciences, Mailman School of Public Health, Columbia University, New York, NY, USA; Columbia Climate School, Columbia University, New York, NY, USA

**Keywords:** COVID-19 forecast, long lead-time, error growth, new variants, infection seasonality

## Abstract

Coronavirus disease 2019 (COVID-19) will likely remain a major public health burden; accurate forecast of COVID-19 epidemic outcomes several months into the future is needed to support more proactive planning. Here, we propose strategies to address three major forecast challenges, i.e., error growth, the emergence of new variants, and infection seasonality. Using these strategies in combination we generate retrospective predictions of COVID-19 cases and deaths 6 months in the future for 10 representative US states. Tallied over >25,000 retrospective predictions through September 2022, the forecast approach using all three strategies consistently outperformed a baseline forecast approach without these strategies across different variant waves and locations, for all forecast targets. Overall, probabilistic forecast accuracy improved by 64% and 38% and point prediction accuracy by 133% and 87% for cases and deaths, respectively. Real-time 6-month lead predictions made in early October 2022 suggested large attack rates in most states but a lower burden of deaths than previous waves during October 2022 – March 2023; these predictions are in general accurate compared to reported data. The superior skill of the forecast methods developed here demonstrate means for generating more accurate long-lead forecast of COVID-19 and possibly other infectious diseases.

**Author Summary:** Infectious disease forecast aims to reliably predict the most likely future outcomes during an epidemic. To date, reliable COVID-19 forecast remains elusive and is needed to support more proactive planning. Here, we pinpoint the major challenges facing COVID-19 forecast and propose three strategies. Comprehensive testing shows the forecast approach using all three strategies consistently outperforms a baseline approach without these strategies across different variant waves and locations in the United States for all forecast targets, improving the probabilistic forecast accuracy by ∼50% and point prediction accuracy by ∼100%. The superior skills of the forecast methods developed here demonstrate means for generating more accurate long-lead COVID-19 forecasts. The methods may be also applicable to other infectious diseases.

**One sentence summary:** To support more proactive planning, we develop COVID-19 forecast methods that substantially improve accuracy with lead time up to 6 months.

## INTRODUCTION

The severe acute respiratory syndrome coronavirus 2 (SARS-CoV-2) emerged in late 2019, causing the coronavirus disease 2019 (COVID-19) pandemic. Since its onset, mathematical modeling has been widely applied to generate projections of potential pandemic trajectories, including for cases, hospitalizations, and deaths. These model projections are often based on specific assumptions (i.e., scenarios) regarding critical factors affecting transmission dynamics. For example, the scenarios often include a combination of public health policies [e.g., non-pharmaceutical interventions (NPIs) including lockdown/reopening and masking, and vaccinations], population behavior (e.g., adherence to the policies and voluntary preventive measures), and anticipated changes in the epidemiological properties of SARS-CoV-2 variants (1–4). While such efforts have provided overviews of the potential outcomes under various scenarios, they do not assign likelihoods to the scenarios/projected trajectories, and the most likely trajectory is typically not known until the outcome is observed. That is, scenario projection is not equivalent to calibrated infectious disease *forecast*, which aims to reliably predict the most likely future outcomes during an epidemic. As COVID-19 will likely remain a major public health burden in the years to come, sensible forecast of the health outcomes several months in the future is needed to support more proactive planning.

Compared to forecast of epidemic infections (e.g., influenza), a number of additional challenges exist for long-lead COVID-19 forecast. First, SARS-CoV-2 new variants will likely continue to emerge and remain a major source of uncertainty when generating COVID-19 forecasts (5, 6). As has been observed for the major variants of concern (VOCs) reported to date (i.e., Alpha, Beta, Gamma, Delta, and Omicron), future new variants could arise at any time, could quickly displace other circulating variants, and could be more contagious than pre-existing variants, and/or erode prior infection- and/or vaccination-induced immunity to affect underlying population susceptibility. Further, multiple new variants could arise successively to cause multiple waves during a time span of, e.g., 6 months. Such frequent emergence and fast turnover of circulating variants is in stark contrast with epidemic infections. Second, many infections for other respiratory viruses tend to occur during a certain season of the year and as such, the seasonality can be incorporated to improve forecast accuracy (7, 8) as well as restrict the forecast window to the epidemic season (e.g., influenza during the winter in temperate regions). Potential seasonality for SARS-CoV-2 is still not well characterized. For instance, in the US, where larger waves have occurred in the winter during 2020-2022, smaller summer waves have also occurred (e.g. the initial Delta wave and Omicron subvariant waves; Fig 1). Third, given the unknown timing of new variant emergence, year-round, long-lead COVID-19 forecast will likely be needed. These unknowns also necessitate wider parameter ranges using a forecast ensemble to account for the uncertainty, which over long forecast horizons could lead to greater error growth and poorer predictive accuracy.

**Fig 1.**
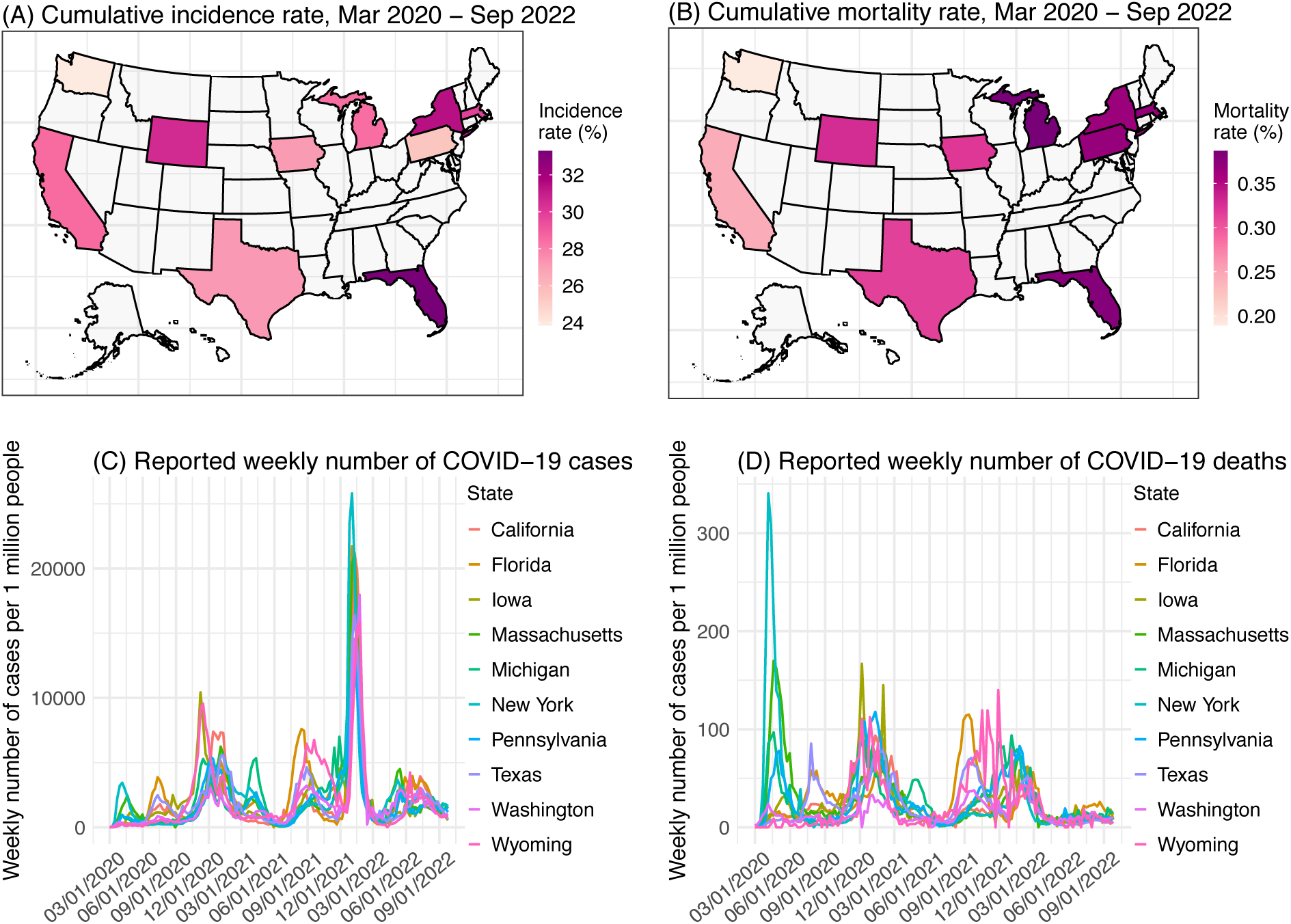
Geospatial distribution of the 10 states and overall COVID-19 outcomes. Heatmaps show reported cumulative COVID-19 incidence rates (A) and COVID-19-associated mortality rates (B) in the 10 states included in this study. Line plots show reported weekly number of COVID-19 cases (C) and COVID-19-associated deaths (D) during the study period, for each state.

In this study, we aim to address the above challenges and develop sensible approaches that support long-lead prediction of COVID-19 epidemic outcomes. We propose three strategies for improving forecast accuracy and test the methods in combination by generating retrospective forecasts of COVID-19 cases and deaths in 10 representative states in the US (i.e., California, Florida, Iowa, Massachusetts, Michigan, New York, Pennsylvania, Texas, Washington, and Wyoming; Fig 1). Relative to a baseline approach, the forecast approach based on our strategies largely improves forecast accuracy (64%/38% higher probabilistic log score for cases/deaths, and 133%/87% higher point prediction accuracy for cases/deaths, tallied over 25,183 evaluations of forecasts initiated during July 2020 – September 2022, i.e., from the end of the initial wave to the time of this study). These results highlight strategies for developing and operationalizing long-lead COVID-19 forecasts with greater demonstrated accuracy and reliability. In addition, we generate real-time COVID-19 forecasts for October 2022 – March 2023 (roughly covering the 2022-2023 respiratory virus season) and evaluate these forecasts using data reported 6 months later.

## RESULTS

### Proposed strategies for long-lead COVID-19 forecast

To address the three challenges noted above, we propose three strategies in combination. Details are provided in Methods. Here, we summarize the idea behind each strategy. The first strategy is to constrain error growth during the extended forecast period. As noted above, the multiple uncertainties regarding SARS-CoV-2 (e.g., new variant properties) necessitate wider distributions of the state variables (e.g., population susceptibility) and parameters (e.g., virus transmissibility) at the point of forecast initiation. With a wider forecast ensemble, some ensemble members could predict earlier, large outbreaks, which if premature, would deplete modeled susceptibility and incorrectly preclude outbreaks later on. More generally, like other infections, COVID-19 epidemics often grow exponentially at first, triggering exponential changes in susceptibility and other state variables, which in turn feed back on the longer epidemic trajectory. Such infectious disease dynamics imply forecast error can also grow exponentially, leading to accelerated degradation of forecast accuracy after the first few weeks. To counteract this exponential error growth, we propose to apply a multiplicative factor γ <1 to the covariance of the forecast state variables (e.g., susceptibility) while retaining the ensemble mean (see Methods). In so doing, we can represent initial uncertainty with a broad forecast ensemble while countering excessive error growth of the state variables. This strategy is similar to covariance inflation during system optimization (9, 10), where a γ >1 is applied to the covariance to counteract over-narrowing of the model ensemble. Since here it acts in the opposite direction, we refer to this technique as deflation. Fig 2A shows example forecasts with deflation compared to without it.

**Fig 2.**
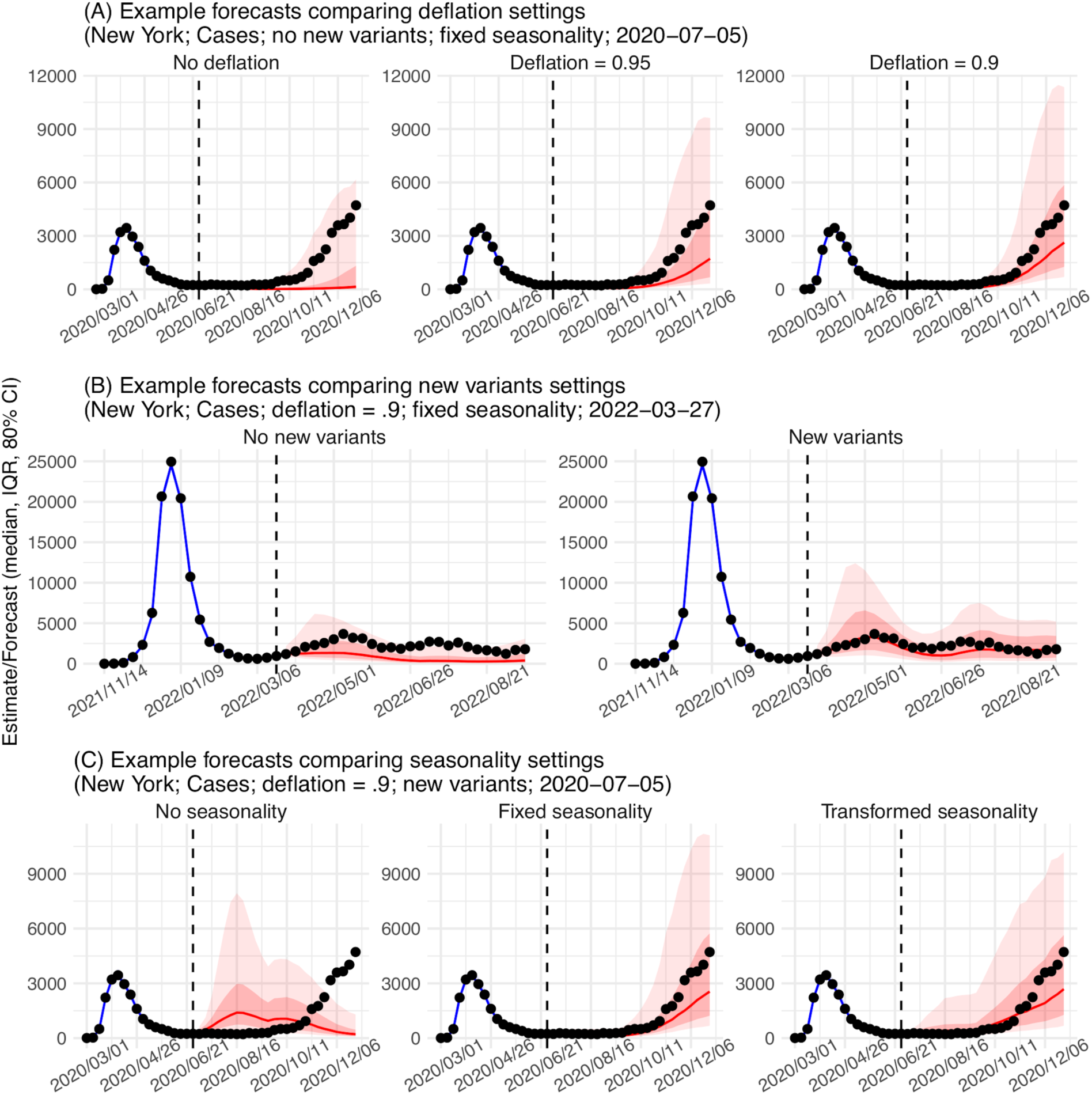
Example forecasts. Vertical dashed lines indicate the week of forecast. Dots show reported weekly cases per 1 million people; only those to the left of the vertical lines are used to calibrate the model and those to the right of the vertical lines are plotted for comparison. Blue lines and blue areas (line = median; darker blue = 50% CI; lighter blue = 80% CI) show model training estimates. Red lines and red areas (line = median; dark red = 50% CI; lighter red = 80% CI) show model forecasts using model settings as labeled in the subtitles.

The second strategy is to anticipate the impact of new variants. Genomic sequencing data can support prediction of the impact of a new variant a few weeks in the future (see Methods and the SI); however, variant displacement and competition dynamics can occur unexpectedly beyond those first few weeks, rendering historical data less relevant. Thus, for weeks farther in the future, instead of predicting specific new variants, we propose to use a set of heuristic rules to anticipate their likely emergence timing and impact on population susceptibility and virus transmissibility. Specifically, for the timing, we reason that new variants are more likely to emerge/circulate 1) after a large local wave when more infections could lead to more mutations, which could be timed based on local outbreak intensity during the preceding months; and 2) during a time when a large part of the world is experiencing a large wave, which also could lead to new mutations. For instance, in the US, this could be during the winter when local large waves tend to occur, or the summer when places in the southern hemisphere are amid their winter waves. As such, this could be timed based on the calendar. For the new variant impact, we observe that, new variant circulation often results in gradual increases in susceptibility (e.g., a few percentages per week, based on estimates from New York City during VOC waves), possibly due to the substantial population immunity accumulated via infections and vaccinations. Similarly, changes in transmissibility also tend to occur gradually. As such, we propose to apply small increases to the model population susceptibility and transmissibility during those two plausible times. The rationale here is to anticipate the more common, non-major changes so as not to overpredict, because major VOCs causing dramatic changes are rarer and difficult to predict. Fig 2B shows example forecasts with these new variant settings compared to without them.

The third strategy accounts for seasonality. Instead of assuming a specific epidemic timing, we model the seasonal risk of SARS-CoV-2 infection based on plausible underlying drivers of infection seasonality for common respiratory viruses. Specifically, studies have shown that respiratory viruses including SARS-CoV-2 are sensitive to ambient humidity and temperature conditions, which could in turn modulate their survival and transmission (11–15). Accordingly, we developed a climate-forced model that includes both humidity and temperature to capture the reported virus response and parameterized the model based on long-term epidemic data observed for influenza (16). When modified and applied to account for SARS-CoV-2 seasonality in a model-inference framework, the estimated seasonal trends are able to capture the effects of different climate conditions (e.g., for UK, Brazil, India, and South Africa (17–19)). Here, we thus propose to apply this model and local climate data to estimate SARS-CoV-2 seasonality in each location (referred to as “fixed seasonality”; see estimates for the 10 states in Fig S1). In addition, as the model is parameterized based on influenza data, this estimated seasonality could differ from the true SARS-CoV-2 seasonality. To test this, we propose an alternative seasonality form that transforms the fixed seasonality trend to allow a more flexible phase timing and structure of seasonality (referred to as “transformed seasonality”; see details in the SI and examples for the 10 states in Fig S1). Fig 2C shows a comparison of the two seasonality forms and example forecasts with no seasonality and the two seasonality forms.

We test the above three strategies in combination, including three deflation settings (i.e., setting γ = 1, 0.95, and 0.9), two new variant settings (i.e., assuming no new variants vs. anticipating new variants per the rules noted above), and three seasonality settings (i.e., no, fixed, and transformed seasonality): in total, 12 (= 3 × 2 × 3) forecast approaches. To compare the performance of the 12 forecast approaches, we generated retrospective forecasts for 10 states, from July 2020 – August 2021 (Pre-Omicron period; 65 weeks in total) and December 2021 – September 2022 (Omicron period; ∼37 weeks). For each week, a forecast for the following 26 weeks (∼6 months) is generated after model training using data up to the week of forecast. We then evaluate the accuracy predicting the weekly number of cases and deaths during each of the 26 weeks (i.e. 1- to 26-week ahead prediction), as well as the peak timing (i.e. the week with the highest cases/deaths), peak intensity, cumulative number of cases and deaths over the 26 weeks.

For the forecast comparisons below, we evaluated probabilistic forecast accuracy using log score, i.e., the logarithm of the probability correctly assigned to the true target (see Methods and SI). We also evaluated point prediction accuracy – assigning value 1 (i.e., accurate) to a forecast if the point prediction is within ±25% of the observed case/death count or within ±1 week of the observed peak week, and 0 (i.e., inaccurate) otherwise; as such, when averaged over all forecasts, this accuracy gives the percentage of forecasts a point prediction is accurate within these tolerances. Thus, both higher log score and higher point prediction accuracy indicate superior forecast performance.

### The impact of deflation

Compared to forecasts with no deflation (*γ* = 1), applying a small deflation (*γ* = 0.95 or 0.9) consistently improves forecast performance (Fig 3, first two panels). This improvement is evident across all locations and combinations of the other two model settings (i.e., new variants and seasonality). The improvement is most pronounced for the long-lead (i.e., 17- to 26-week ahead) weekly forecasts and overall intensity-related targets (i.e., the totals accumulated over 26 forecast weeks and peak intensity; Figs S2–3), indicating deflation is able to effectively reduce error growth accumulated over time. We note that, as deflation works by reducing forecast spread, the forecast ensemble can also become under-dispersed, assign zero probability to the true target (most notably, for peak week), and in turn produce a lower log score (see Fig S2, the 3^rd^ row of each heatmap for peak week). Given this trade-off, we did not test deflation factors <0.9.

**Fig 3.**
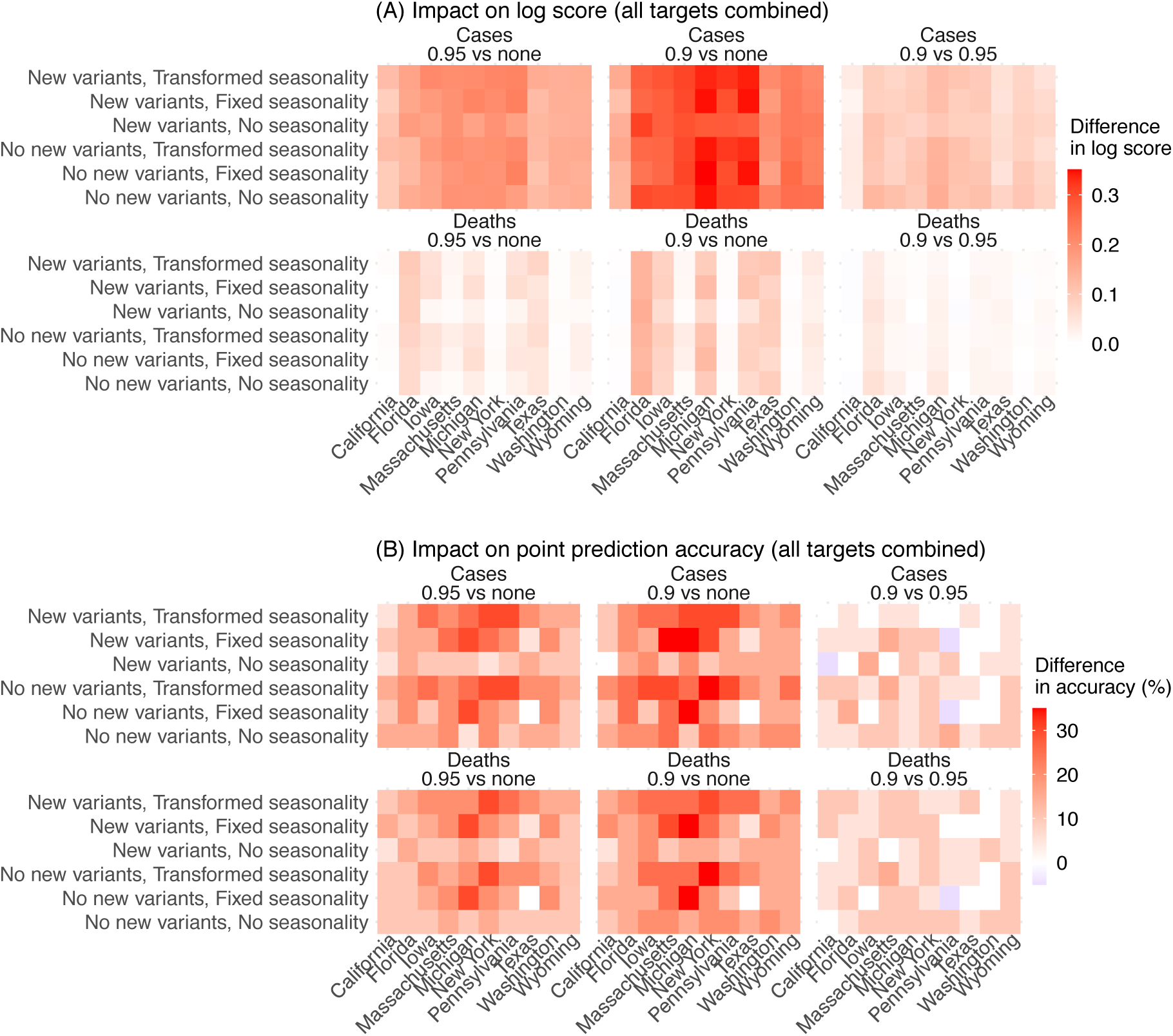
Impact of deflation on forecast performance. Heatmaps show the differences in mean log score (A) or point prediction accuracy (B) between all forecast approaches with different deflation settings (deflation factor γ = 0.95 vs none in the 1^st^ column, 0.9 vs none in the 2^nd^ column, and 0.9 vs 0.95 in the 3^rd^ column; see panel subtitles). Results are aggregated for each forecast approach (see specific settings of new variants and seasonality in the y-axis labels) and location (x-axis) over all forecast targets and forecast weeks, for cases (1^st^ row) and deaths (2^nd^ row), separately. For each pairwise comparison (e.g., 0.95 vs none), a positive difference in log score or point prediction accuracy indicates the former approach (e.g., 0.95) outperforms the latter (e.g., none).

Overall, relative to forecasts without deflation (*γ* = 1), log scores aggregated over all locations and targets were 18 – 20% higher for forecasts of cases (range of relative change across all combinations of the other two model settings; Table S1) and 7 – 8% higher for forecasts of mortality when a deflation factor *γ* of 0.95 was applied. The log scores further increased when using *γ* = 0.9, to 34 – 43% higher (than *γ* = 1) for forecasts of cases and 13 – 17% higher for forecasts of mortality. The improvement of point prediction accuracy was more pronounced. Aggregated over all locations and targets, point prediction accuracy was 33 – 63% higher for forecasts of cases and 24 – 40% higher for forecasts of mortality when using a deflation factor *γ* of 0.9, relative to using no deflation (Table S1). As such, we use *γ* = 0.9 as the best-performing setting for subsequent analyses.

### Impact of the new variant settings

To ensure consistency and avoid over-fitting, we applied the same set of heuristic rules on new variant emergence timing and impact, as noted above, throughout the entire study period (i.e., including weeks before the emergence of SARS-CoV-2 VOCs). We expect the new variant settings to improve forecast performance during VOC waves but have a less pronounced or no effect during the 2^nd^ wave (roughly, fall/winter 2020 – 2021), prior to VOC emergence. Indeed, overall, for the 2^nd^ wave, forecast systems with the new variant settings (referred to as “new variant model”) had similar performance as those assuming no new variants (relative changes in log score and accuracy: −2% to 0.6%, Table S2; Fig 4). However, for the VOC waves, applying the new variant settings improved forecast performance during the Alpha wave (roughly, spring 2021), Delta wave (roughly, summer/fall 2021), and Omicron wave (after December 2021; note no further desegregation was made for Omicron subvariant waves due to small sample sizes; Fig 4). For forecasts of cases, the improvement was consistently seen for all three VOC waves (relative changes in log score and accuracy all > 0, Table S2). The relative increases of log score were up to 119% during the Delta wave and up to 37% during the Omicron wave; the relative increases of point prediction accuracies were up to 89% during the Delta wave and up to 96% during the Omicron wave (Table S2 and Fig 4).

**Fig 4.**
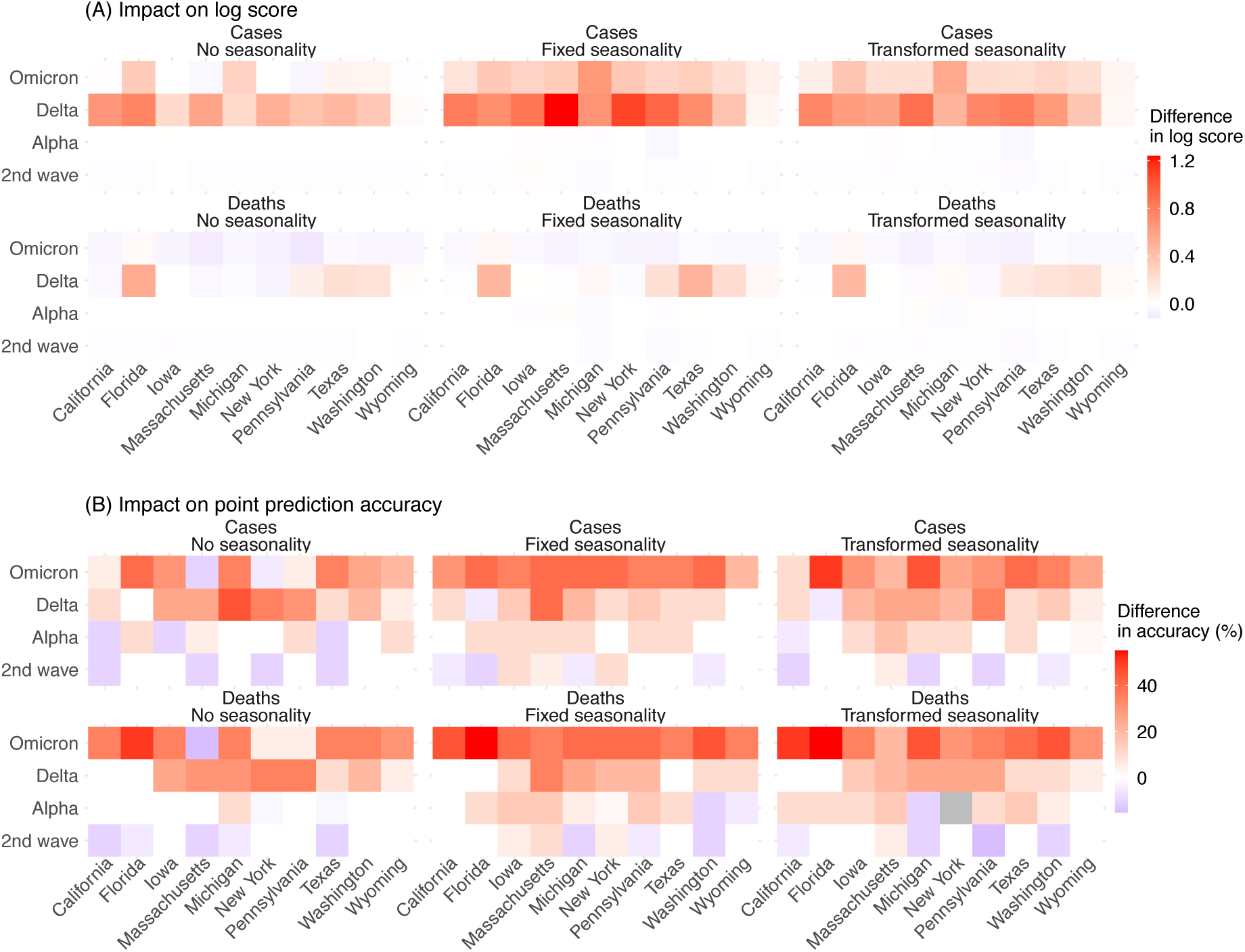
Impact of new variant settings on forecast performance. Heatmaps show the differences in mean log score (A) or point prediction accuracy (B) between forecast approaches with vs without anticipation of new variant emergence. All forecasts here were generated using a deflation factor of 0.9. Results are aggregated for each forecast approach (see specific setting of seasonality in panel subtitles), variant wave (y-axis), and location (x-axis) over all forecast targets and forecast weeks for cases (1^st^ row) and deaths (2^nd^ row), separately. A positive difference indicates superior performance of the forecast approach with anticipation of new variant emergence.

For forecasts of mortality, the new variant model had higher log scores during the Alpha and Delta waves, as well as higher point prediction accuracies during all three VOC waves (Table S2). However, log scores during the Omicron wave were similar for both models (e.g., overall log scores: −0.22 for the new variant model and −0.17 for the baseline, both assuming no seasonality); in addition, the log scores were slightly lower for the new variant model, likely due to the lower COVID-19-associated deaths during the Omicron wave.

Aggregated over all waves, targets, and locations, the new variant model had 17 – 34% and 3 – 8% higher log scores and 23 – 28% and 22 – 25% higher point prediction accuracies for forecasts of cases and deaths, respectively.

### Impact of seasonality forms

Based on the above results, we focus on forecasts generated using the new variant model with a deflation factor of 0.9 to examine the three approaches to forecasting the effects of seasonality. As noted above, seasonality aims to capture changes in infection risk in response to environmental conditions (here, ambient humidity and temperature); for common respiratory viruses (e.g., influenza), infection risk in temperate regions is often higher during cold-dry winter months (i.e., respiratory virus season, roughly mid-October to mid-April in the US), and lower during the rest of the year (i.e., off season). Both the fixed and transformed seasonality models consistently improved forecast performance during the respiratory virus season relative to the no seasonality approach across all locations and targets (Fig 5A for log score and 5B for point prediction accuracy, first two panels; Table S3). However, if only analyzing the off season, both models had worse performance compared to the model assuming no seasonality (Fig 5, 4^th^ and 5^th^ panels). Segregating the forecasts made for the off season by wave shows that both seasonality models continued to outperform the no seasonality model during summer/fall 2020 (grouped with the 2^nd^ wave); worse performance occurred during summer/fall 2021 (the Alpha and Delta waves) and summer 2022 (Omicron subvariants; Table S3). These results suggest that the seasonality models are able to capture the seasonal risk of SARS-CoV-2 infection, and that the degraded performance during the off season may be due to challenges anticipating the initial surge of VOCs occurring during those times.

**Fig 5.**
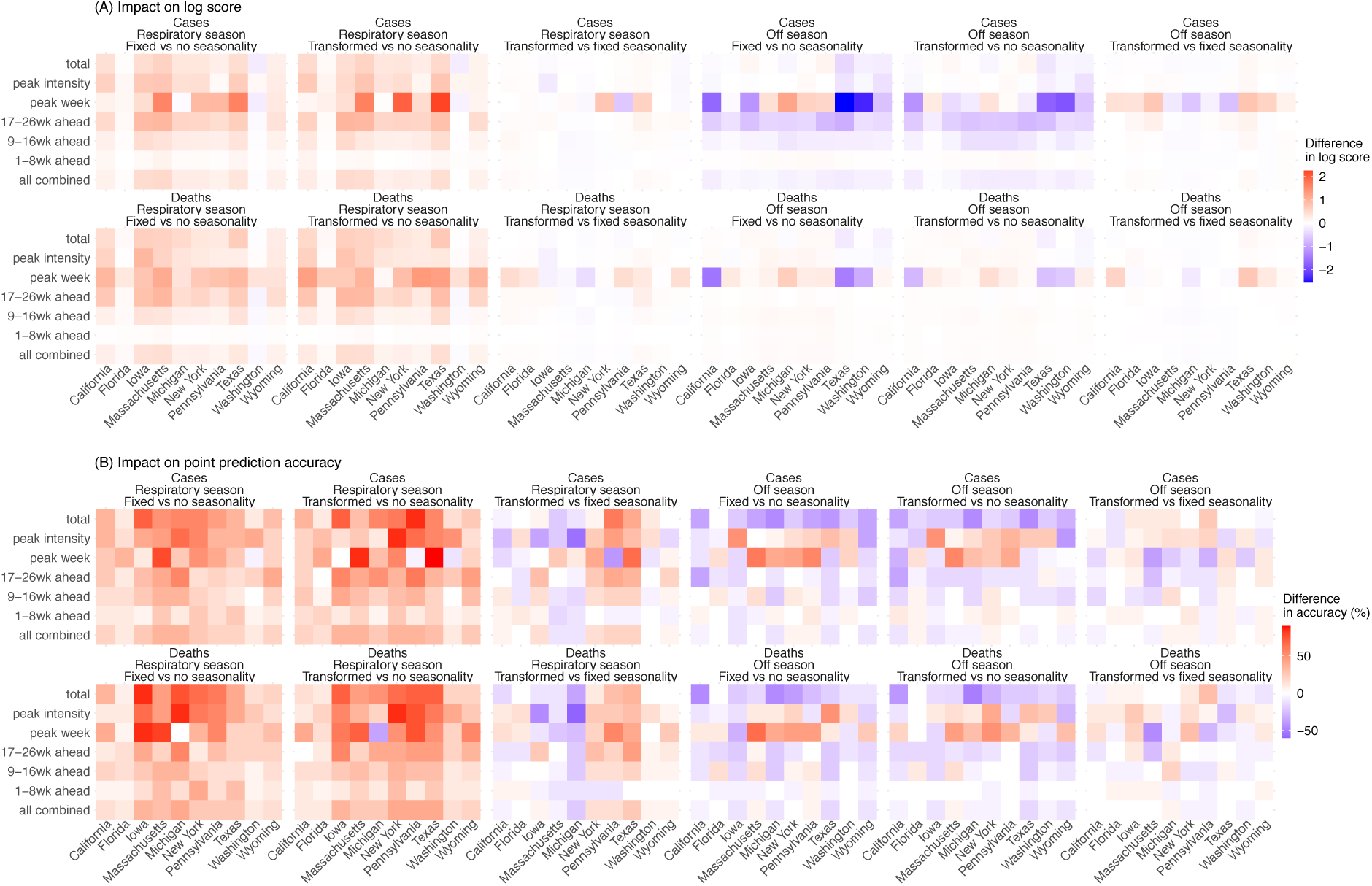
Impact of seasonality settings on forecast performance. Heatmaps show the differences in mean log score (A) or point prediction accuracy (B), between pairs of forecast approaches with different seasonality settings (see panel subtitles). All forecasts here were generated using a deflation factor of 0.9 and the new variant setting. Results are aggregated for each forecast target (y-axis) and location (x-axis), over either the respiratory virus season (first 3 columns) or the off season (last 3 columns), for cases (1^st^ row) and deaths (2^nd^ row), separately. For each pairwise comparison (e.g., fixed vs no seasonality), a positive difference in log score or point prediction accuracy indicates the former approach (e.g., with fixed seasonality) outperforms the latter (e.g., with no seasonality).

Tallied over all time periods, in general, the seasonality models outperformed the no seasonality model (Table S3 for all locations combined and Table S4 for each location). Comparing the two seasonality forms, the transformed seasonality model outperformed the fixed seasonality model overall (Table S3). Compared to the no seasonality model, the transformed seasonality had 14% higher log score for forecasts of mortality, and 26% and 18% higher point prediction accuracies for cases and mortality, respectively (Table S3). As noted above, the improvement during the respiratory virus season was more substantial (Tables S3 and S4; Fig 5).

### Combined impact of deflation, new variant settings, and seasonality forms

We now examine the forecast performance using the combined best-performing approach (i.e., applying deflation with γ = 0.9, the new variant rules, and the transformed seasonality form), compared to the baseline approach (i.e., no deflation, no new variants, and no seasonality). In addition to the large uncertainties surrounding SARS-CoV-2 (e.g., new variants), there are also large spatial heterogeneities. For example, across the 10 states included here, population density ranged from 6 people per square mile in Wyoming to 884 per square mile in Massachusetts (2020 data (20)); climate conditions span temperate (e.g., New York) and subtropical climates (e.g., Florida; Fig S1). Given the uncertainties and spatial heterogeneities, the robustness of any forecast approach is particularly important.

First, we examine the consistency of forecast performance over different variant periods. During the pre-Omicron period (here, July 2020 – Dec 2021), the combined approach consistently outperformed the baseline approach across all states, for both forecasts of cases and deaths (Fig 6A); log scores improved by 85% overall for cases (range from 39% in Washington to 134% in Florida) and by 62% overall for deaths (range from 24% in Washington to 117% in Florida; Table 1). During the Omicron period (here, Dec 2021 – September 2022), improvements were smaller but consistent across the 10 states for forecast of cases (Fig 6A; note that only 16-20 forecasts of long-lead targets were evaluated here, as observations are incomplete); as noted above, due to the much lower mortality during the Omicron period, both the best-performing and baseline forecasts of mortality had similar log scores (e.g., median difference = −0.02, Table 1). The overall consistency of the performance indicates that the best-performing forecast approach is robust for forecasting long-lead COVID-19 epidemic outcomes for different variants.

**Fig 6.**
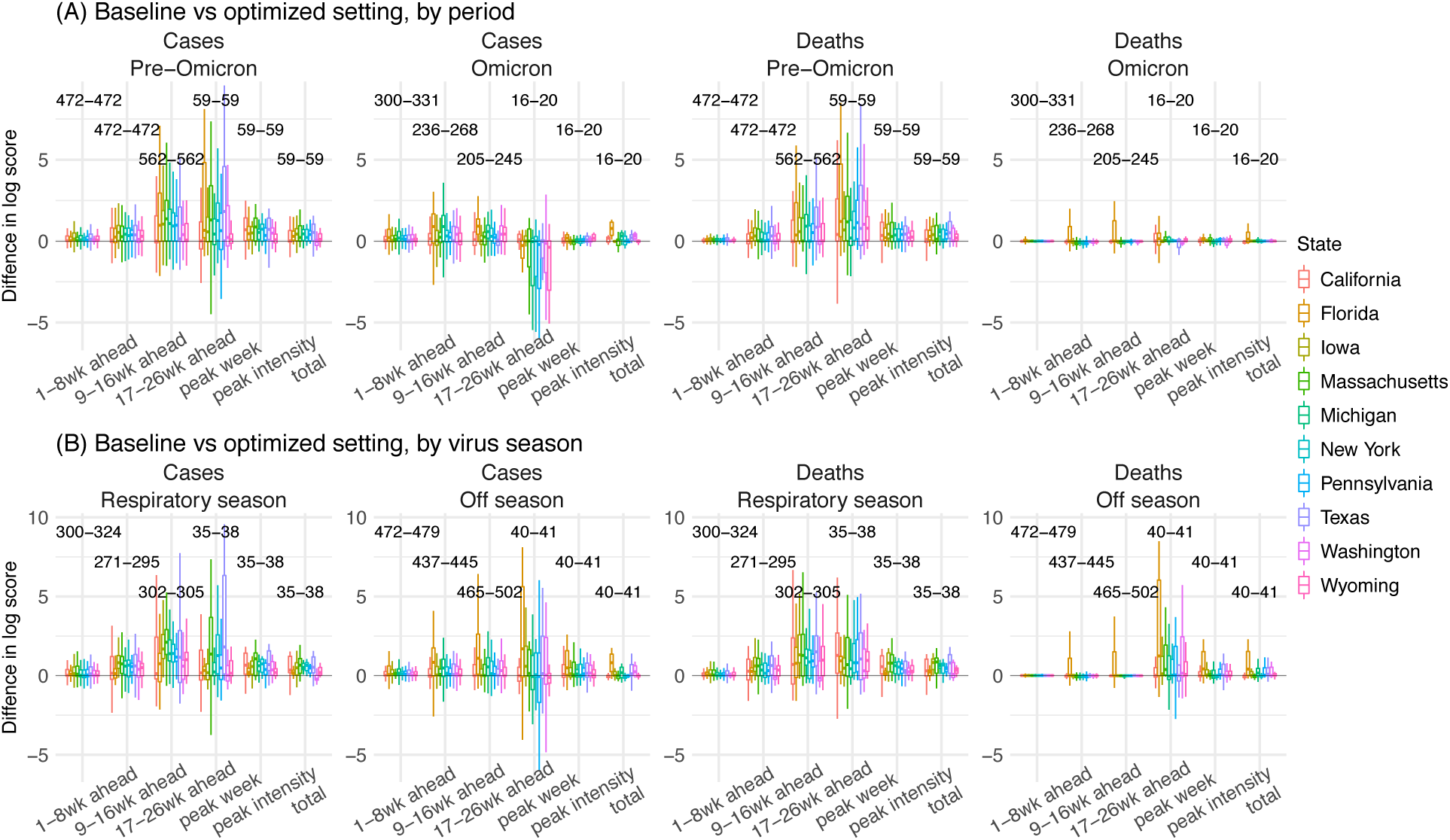
Probabilistic forecast accuracy of the best-performing and baseline forecast approaches. Boxplots show the distributions of pair-wise difference in log score by variant period (A) or respiratory virus season (B; see panel subtitles). Results are aggregated by location (color-coded for each state) and forecast target (x-axis), for cases and deaths (see panel subtitles), separately. The numbers show the range of number of evaluations of each forecast target (e.g., 59 predictions of peak week during the pre-Omicron period, for each state; 16-20 predictions of peak week during the Omicron period, depending on the timing of Omicron detection in each state). A positive difference indicates superior log score of the best-performing forecast approach.

**Table 1.**
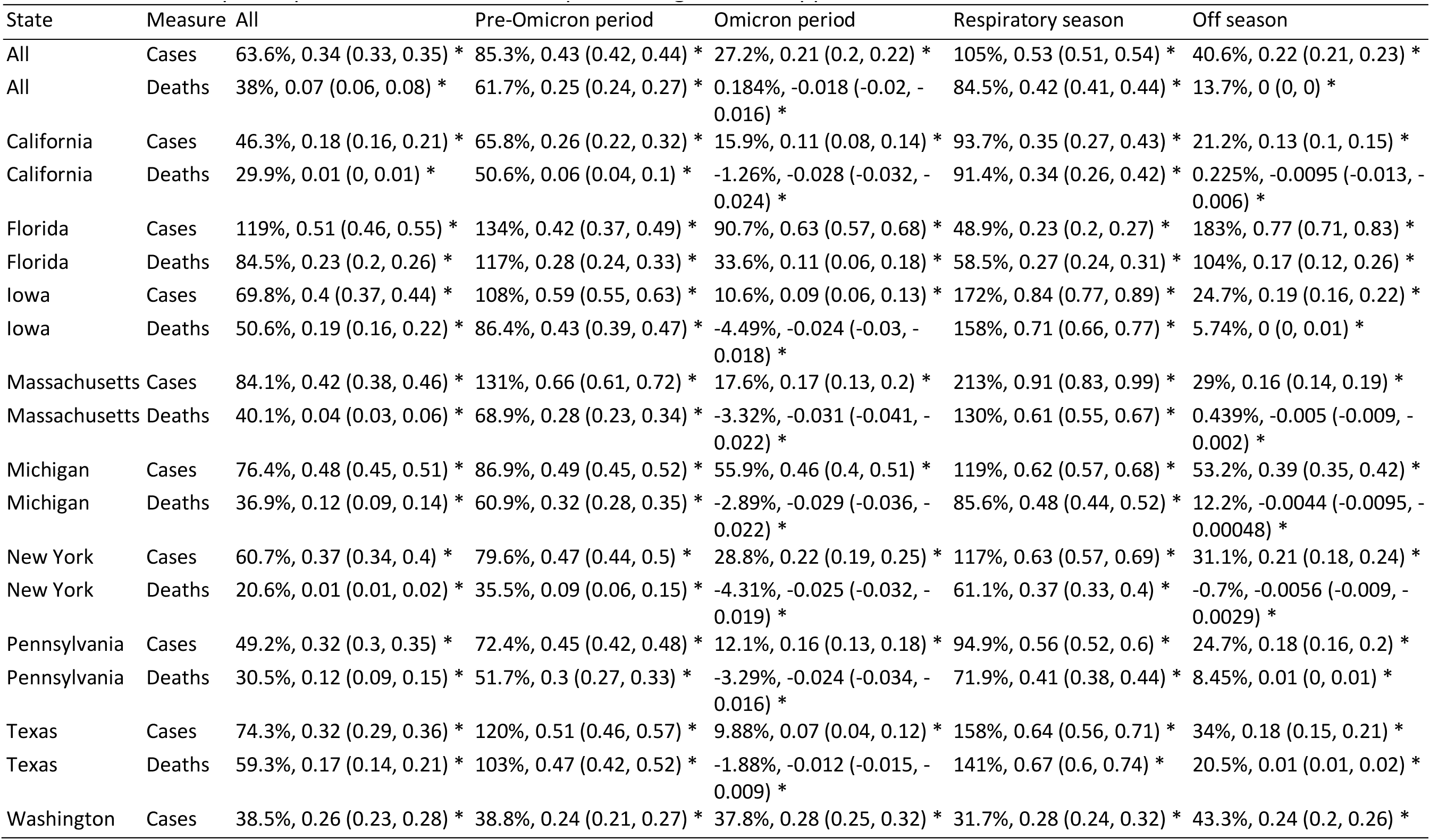
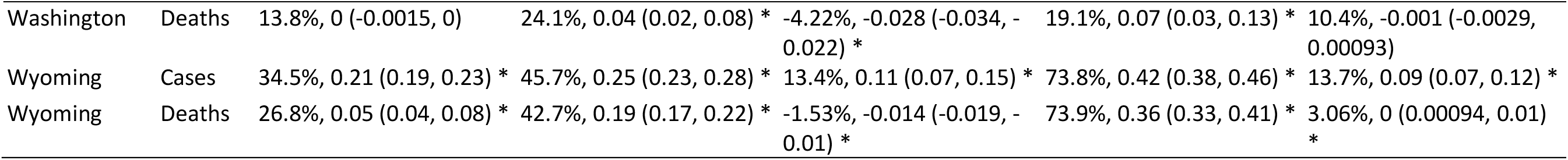
Comparison of probabilistic forecast accuracy by the best-performing and the baseline forecast approaches. Numbers show the relative difference in mean log score computed using Eqn 6, the median of pairwise difference in log score (95% CI of the median); asterisk (*) indicates if the median is significantly >0 or <0 at the α = 0.05 level, per a Wilcoxon rank sum test. Positive numbers indicate superior performance of the best-performing forecast approach.

| State | Measure | All | Pre-Omicron period | Omicron period | Respiratory season | Off season |
| --- | --- | --- | --- | --- | --- | --- |
| All | Cases | 63.6%, 0.34 (0.33, 0.35) * | 85.3%, 0.43 (0.42, 0.44) * | 27.2%, 0.21 (0.2, 0.22) * | 105%, 0.53 (0.51, 0.54) * | 40.6%, 0.22 (0.21, 0.23) * |
| All | Deaths | 38%, 0.07 (0.06, 0.08) * | 61.7%, 0.25 (0.24, 0.27) * | 0.184%, -0.018 (-0.02, -0.016) * | 84.5%, 0.42 (0.41, 0.44) * | 13.7%, 0 (0, 0) * |
| California | Cases | 46.3%, 0.18 (0.16, 0.21) * | 65.8%, 0.26 (0.22, 0.32) * | 15.9%, 0.11 (0.08, 0.14) * | 93.7%, 0.35 (0.27, 0.43) * | 21.2%, 0.13 (0.1, 0.15) * |
| California | Deaths | 29.9%, 0.01 (0, 0.01) * | 50.6%, 0.06 (0.04, 0.1) * | -1.26%, -0.028 (-0.032, -0.024) * | 91.4%, 0.34 (0.26, 0.42) * | 0.225%, -0.0095 (-0.013, -0.006) * |
| Florida | Cases | 119%, 0.51 (0.46, 0.55) * | 134%, 0.42 (0.37, 0.49) * | 90.7%, 0.63 (0.57, 0.68) * | 48.9%, 0.23 (0.2, 0.27) * | 183%, 0.77 (0.71, 0.83) * |
| Florida | Deaths | 84.5%, 0.23 (0.2, 0.26) * | 117%, 0.28 (0.24, 0.33) * | 33.6%, 0.11 (0.06, 0.18) * | 58.5%, 0.27 (0.24, 0.31) * | 104%, 0.17 (0.12, 0.26) * |
| Iowa | Cases | 69.8%, 0.4 (0.37, 0.44) * | 108%, 0.59 (0.55, 0.63) * | 10.6%, 0.09 (0.06, 0.13) * | 172%, 0.84 (0.77, 0.89) * | 24.7%, 0.19 (0.16, 0.22) * |
| Iowa | Deaths | 50.6%, 0.19 (0.16, 0.22) * | 86.4%, 0.43 (0.39, 0.47) * | -4.49%, -0.024 (-0.03, -0.018) * | 158%, 0.71 (0.66, 0.77) * | 5.74%, 0 (0, 0.01) * |
| Massachusetts | Cases | 84.1%, 0.42 (0.38, 0.46) * | 131%, 0.66 (0.61, 0.72) * | 17.6%, 0.17 (0.13, 0.2) * | 213%, 0.91 (0.83, 0.99) * | 29%, 0.16 (0.14, 0.19) * |
| Massachusetts | Deaths | 40.1%, 0.04 (0.03, 0.06) * | 68.9%, 0.28 (0.23, 0.34) * | -3.32%, -0.031 (-0.041, -0.022) * | 130%, 0.61 (0.55, 0.67) * | 0.439%, -0.005 (-0.009, -0.002) * |
| Michigan | Cases | 76.4%, 0.48 (0.45, 0.51) * | 86.9%, 0.49 (0.45, 0.52) * | 55.9%, 0.46 (0.4, 0.51) * | 119%, 0.62 (0.57, 0.68) * | 53.2%, 0.39 (0.35, 0.42) * |
| Michigan | Deaths | 36.9%, 0.12 (0.09, 0.14) * | 60.9%, 0.32 (0.28, 0.35) * | -2.89%, -0.029 (-0.036, -0.022) * | 85.6%, 0.48 (0.44, 0.52) * | 12.2%, -0.0044 (-0.0095, -0.00048) * |
| New York | Cases | 60.7%, 0.37 (0.34, 0.4) * | 79.6%, 0.47 (0.44, 0.5) * | 28.8%, 0.22 (0.19, 0.25) * | 117%, 0.63 (0.57, 0.69) * | 31.1%, 0.21 (0.18, 0.24) * |
| New York | Deaths | 20.6%, 0.01 (0.01, 0.02) * | 35.5%, 0.09 (0.06, 0.15) * | -4.31%, -0.025 (-0.032, -0.019) * | 61.1%, 0.37 (0.33, 0.4) * | -0.7%, -0.0056 (-0.009, -0.0029) * |
| Pennsylvania | Cases | 49.2%, 0.32 (0.3, 0.35) * | 72.4%, 0.45 (0.42, 0.48) * | 12.1%, 0.16 (0.13, 0.18) * | 94.9%, 0.56 (0.52, 0.6) * | 24.7%, 0.18 (0.16, 0.2) * |
| Pennsylvania | Deaths | 30.5%, 0.12 (0.09, 0.15) * | 51.7%, 0.3 (0.27, 0.33) * | -3.29%, -0.024 (-0.034, -0.016) * | 71.9%, 0.41 (0.38, 0.44) * | 8.45%, 0.01 (0, 0.01) * |
| Texas | Cases | 74.3%, 0.32 (0.29, 0.36) * | 120%, 0.51 (0.46, 0.57) * | 9.88%, 0.07 (0.04, 0.12) * | 158%, 0.64 (0.56, 0.71) * | 34%, 0.18 (0.15, 0.21) * |
| Texas | Deaths | 59.3%, 0.17 (0.14, 0.21) * | 103%, 0.47 (0.42, 0.52) * | -1.88%, -0.012 (-0.015, -0.009) * | 141%, 0.67 (0.6, 0.74) * | 20.5%, 0.01 (0.01, 0.02) * |
| Washington | Cases | 38.5%, 0.26 (0.23, 0.28) * | 38.8%, 0.24 (0.21, 0.27) * | 37.8%, 0.28 (0.25, 0.32) * | 31.7%, 0.28 (0.24, 0.32) * | 43.3%, 0.24 (0.2, 0.26) * |
| Washington | Deaths | 13.8%, 0 (-0.0015, 0) | 24.1%, 0.04 (0.02, 0.08) * | -4.22%, -0.028 (-0.034, -0.022) * | 19.1%, 0.07 (0.03, 0.13) * | 10.4%, -0.001 (-0.0029, 0.00093) |
| Wyoming | Cases | 34.5%, 0.21 (0.19, 0.23) * | 45.7%, 0.25 (0.23, 0.28) * | 13.4%, 0.11 (0.07, 0.15) * | 73.8%, 0.42 (0.38, 0.46) * | 13.7%, 0.09 (0.07, 0.12) * |
| Wyoming | Deaths | 26.8%, 0.05 (0.04, 0.08) * | 42.7%, 0.19 (0.17, 0.22) * | -1.53%, -0.014 (-0.019, -0.01) * | 73.9%, 0.36 (0.33, 0.41) * | 3.06%, 0 (0.00094, 0.01) * |

Second, we examine the forecast performance during the US respiratory virus season (here, mid-October to mid-April) when larger COVID-19 waves have occurred. Tallied over all weeks during the respiratory virus season, the best-performing approach outperformed the baseline approach for all 10 states (Fig 6B); log scores increased by 105% for cases (range from 32% in Washington to 213% in Massachusetts) and by 85% for mortality (range from 19% in Washington to 158% in Iowa; Table 1). During the off season, the best-performing approach also generally outperformed the baseline approach (Table 1).

Third, we examine the accuracy predicting different epidemic targets. The best-performing forecast approach consistently improved point prediction accuracy for all targets for all 10 states (Fig 7 and Table 2). In addition, the improvement was more substantial for long-lead targets (e.g., 9- to 16-week ahead and 17- to 26-week ahead forecasts, peak week, peak intensity, and the cumulative totals). For instance, the best-performing approach increased accuracy from 24% to 42% (35% to 56%) predicting the peak week of cases (deaths), from 20% to 40% (30% to 50%) predicting the peak intensity of cases (deaths), roughly a 2-fold improvement for these two long-lead targets. The improvements were even more substantial for 17-26 week ahead forecasts and the cumulative totals over the entire 26 weeks (by 3- to 22-fold, Fig 7 and Table 2). We note the forecasts here were generated retrospectively with information that may not be available in real time and thus likely are more accurate as a result. Nonetheless, with the same information provided to both forecast approaches, the comparison here demonstrates the large improvement in forecast accuracy using the best-performing forecast approach.

**Fig 7.**
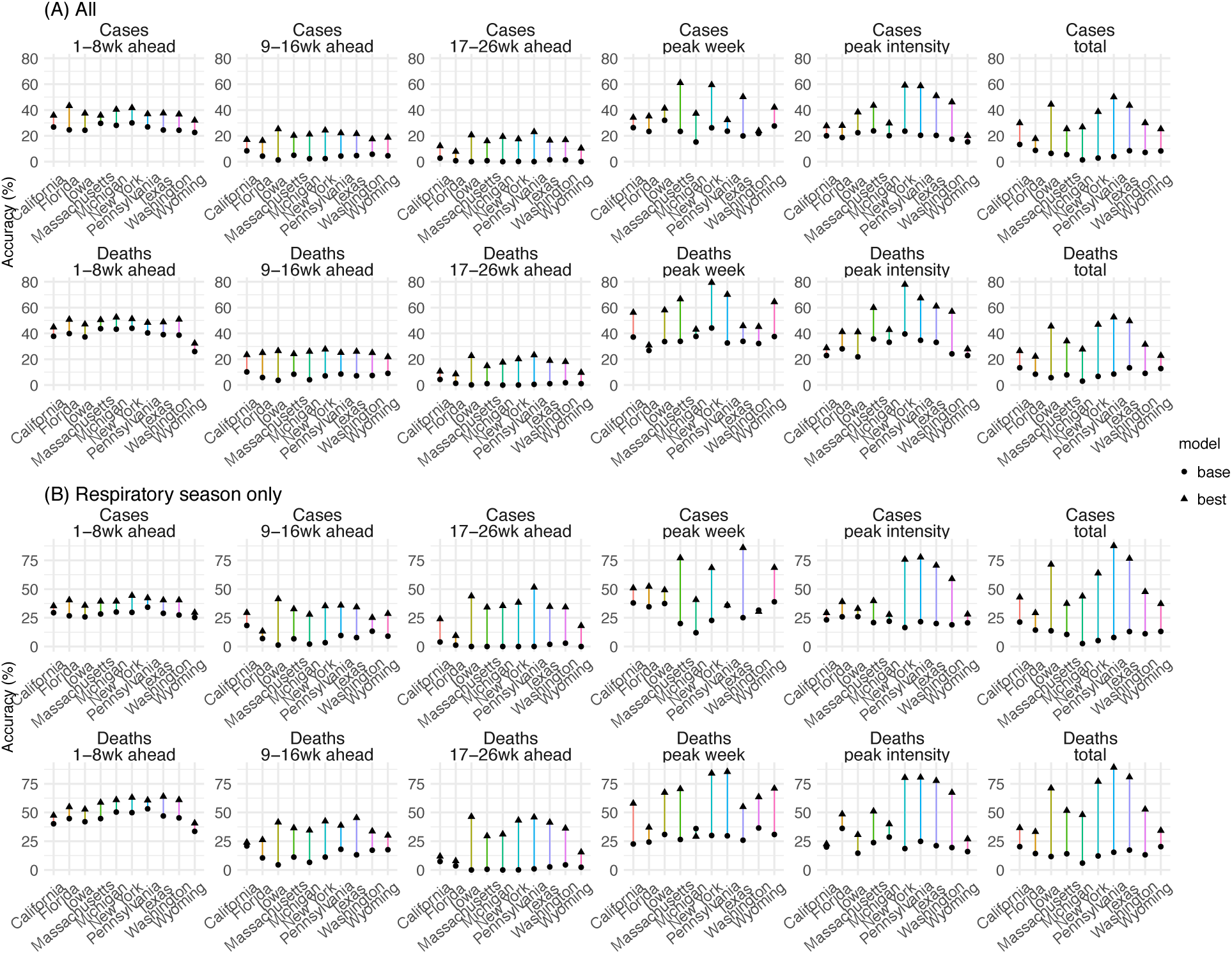
Point prediction accuracy of the best-performing and baseline forecast systems. Points show the average accuracy over all forecast weeks (A) or respiratory virus season (B). Results are aggregated by location (x-axis) and forecast target (panel subtitles) for cases (1^st^ row) and deaths (2^nd^ row, see panel subtitles) separately. Filled dots show the mean accuracy of forecasts generated using the baseline system; filled triangles show the accuracy of forecasts generated using the best-performing forecast system. The lines linking the two accuracies show the changes (mostly increases, as the triangles are more often above the dots), due to the combined application of the three proposed strategies (deflation, new variants, and transformed seasonality settings). Note all forecasts were generated retrospectively; to enable comparison of the model settings, mobility and vaccination data and estimates of infection detection rate and infection fatality risk during the forecast period were used (see main text for detail).

**Table 2.** Comparison of point prediction accuracy by the best-performing and the baseline forecast approaches. Numbers show the mean point prediction accuracy of forecasts generated using the baseline v. the best-performing forecast approach; asterisk (*) indicates if the median of pairwise accuracy difference is significantly >0 or <0 at the α = 0.05 level, per a Wilcoxon rank sum test. Note all forecasts were generated retrospectively; to enable comparison of forecast approaches, mobility and vaccination data and estimates of infection detection rate and infection fatality risk during the forecast period were used (see main text for detail).

| State | Measure | 1-8wk ahead | 9-16wk ahead | 17-26wk ahead | peak week | peak intensity | total |
| --- | --- | --- | --- | --- | --- | --- | --- |
| All | Cases | 26% v 38% * | 4% v 20% * | 1% v 16% * | 24% v 42% * | 20% v 40% * | 7% v 33% * |
| All | Deaths | 39% v 48% * | 7% v 25% * | 1% v 16% * | 35% v 56% * | 30% v 50% * | 9% v 36% * |
| California | Cases | 27% v 36% * | 8% v 17% * | 3% v 12% * | 26% v 34% | 20% v 28% * | 13% v 30% * |
| California | Deaths | 38% v 45% * | 10% v 23% * | 4% v 11% * | 37% v 56% * | 23% v 29% | 13% v 26% * |
| Florida | Cases | 25% v 43% * | 4% v 16% * | 1% v 8% * | 23% v 35% * | 19% v 28% * | 9% v 18% * |
| Florida | Deaths | 40% v 51% * | 6% v 25% * | 1% v 8% * | 27% v 31% | 28% v 41% * | 8% v 22% * |
| Iowa | Cases | 24% v 37% * | 1% v 25% * | 0% v 21% * | 32% v 41% * | 22% v 38% * | 6% v 44% * |
| Iowa | Deaths | 37% v 47% * | 4% v 26% * | 0% v 23% * | 34% v 58% * | 22% v 41% * | 6% v 46% * |
| Massachusetts | Cases | 30% v 36% * | 5% v 20% * | 1% v 16% * | 23% v 61% * | 24% v 43% * | 5% v 25% * |
| Massachusetts | Deaths | 44% v 50% * | 8% v 24% * | 1% v 15% * | 34% v 67% * | 36% v 60% * | 8% v 34% * |
| Michigan | Cases | 28% v 40% * | 2% v 21% * | 0% v 19% * | 15% v 37% * | 20% v 30% * | 1% v 27% * |
| Michigan | Deaths | 43% v 52% * | 4% v 26% * | 0% v 18% * | 38% v 43% | 33% v 43% * | 3% v 28% * |
| New York | Cases | 30% v 42% * | 2% v 24% * | 0% v 18% * | 26% v 59% * | 24% v 59% * | 3% v 39% * |
| New York | Deaths | 44% v 51% * | 7% v 28% * | 0% v 20% * | 44% v 79% * | 40% v 78% * | 7% v 47% * |
| Pennsylvania | Cases | 27% v 37% * | 4% v 22% * | 0% v 23% * | 24% v 32% * | 20% v 59% * | 4% v 50% * |
| Pennsylvania | Deaths | 40% v 48% * | 9% v 25% * | 1% v 23% * | 33% v 70% * | 35% v 67% * | 9% v 53% * |
| Texas | Cases | 24% v 38% * | 5% v 22% * | 1% v 16% * | 20% v 50% * | 20% v 51% * | 8% v 44% * |
| Texas | Deaths | 39% v 49% * | 7% v 26% * | 1% v 19% * | 34% v 46% * | 33% v 61% * | 13% v 50% * |
| Washington | Cases | 24% v 37% * | 6% v 17% * | 1% v 17% * | 22% v 24% | 17% v 46% * | 7% v 30% * |
| Washington | Deaths | 39% v 51% * | 7% v 25% * | 2% v 18% * | 32% v 45% * | 24% v 57% * | 9% v 31% * |
| Wyoming | Cases | 23% v 32% * | 5% v 19% * | 0% v 10% * | 28% v 42% * | 15% v 20% * | 8% v 25% * |
| Wyoming | Deaths | 26% v 32% * | 9% v 22% * | 1% v 10% * | 38% v 64% * | 23% v 28% * | 13% v 23% * |

### Forecast performance compared with ARIMAX models

To benchmark the performance of the forecast approaches developed here, we also generated retrospective forecasts using Auto-Regressive Integrated Moving Average (ARIMA) models. Compared to the best-performing ARIMAX model (identified from 5 models with different settings; see Methods and Table S5), our baseline approach (i.e., no deflation, no new variants, and no seasonality) performed similarly well whereas our best-performing approach (i.e., applying deflation with γ = 0.9, the new variant rules, and the transformed seasonality form) had much superior performance (Table S6).

### Forecast for the 2022 – 2023 respiratory virus season

Figs 8–9 present real-time forecasts of October 2022 – March 2023 for the 10 states, and Table S7 shows a preliminary accuracy assessment based on data obtained on March 31, 2023. Accounting for under-detection, large numbers of infections (i.e., including undocumented asymptomatic or mild infections) were predicted in the coming months for most states; predicted attack rates over the 6-month prediction period ranged from 16% (IQR: 7 – 31%) in Florida to 30% (IQR: 15 – 47%) in Massachusetts (Fig 9). Relatively low case numbers and fewer deaths at levels similar to or lower than previous waves were forecast, assuming case ascertainment rates and infection-fatality risks similar to preceding weeks (Fig 9). Compared to data reported 6 months later (i.e., not used in the forecasts), the weekly forecasts in general captured trajectories of reported weekly cases over the 6 months for all 10 states (Fig 8, middle column for each state) but under-predicted deaths for half of the states (i.e., New York, Massachusetts, Michigan, Wyoming, and Florida; Fig 8, right column for each state). For the cumulative totals, predicted IQRs covered reported tallies in all 10 states for cases and the majority of states for deaths, while the 95% predicted intervals covered reported cumulative cases and deaths in all states (Fig 9).

**Fig 8.**
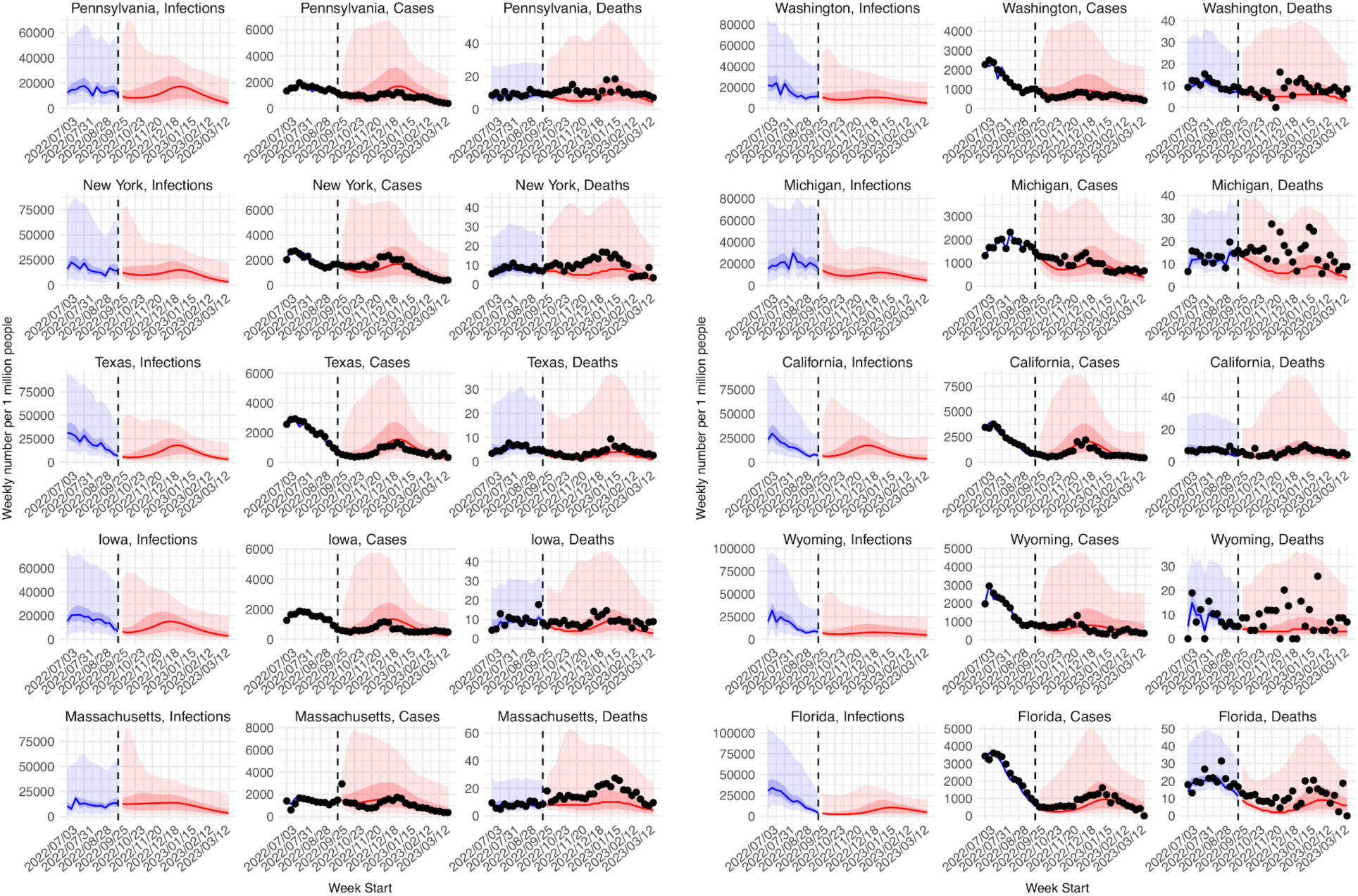
Real-time forecasts for the 2022-2023 respiratory virus season. The states are arranged based on accuracy of historical forecast (higher accuracy for those in the left panel and those on the top). In each panel, each row shows estimates and forecasts of weekly numbers of infections (1^st^ column), cases (2^nd^ column), or deaths (3^rd^ column) for each state. Vertical dashed lines indicate the week of forecast initiation (i.e., October 2, 2022). Dots show reported weekly cases or deaths, including for the forecast period. Blue lines and blue areas (line = median; darker blue = 50% CI; lighter blue = 95% CI) show model training estimates. Red lines and red areas (line = median; dark red = 50% Predictive Interval; lighter red = 95% Predictive Interval) show model forecasts using the best-performing approach.

**Fig 9.**
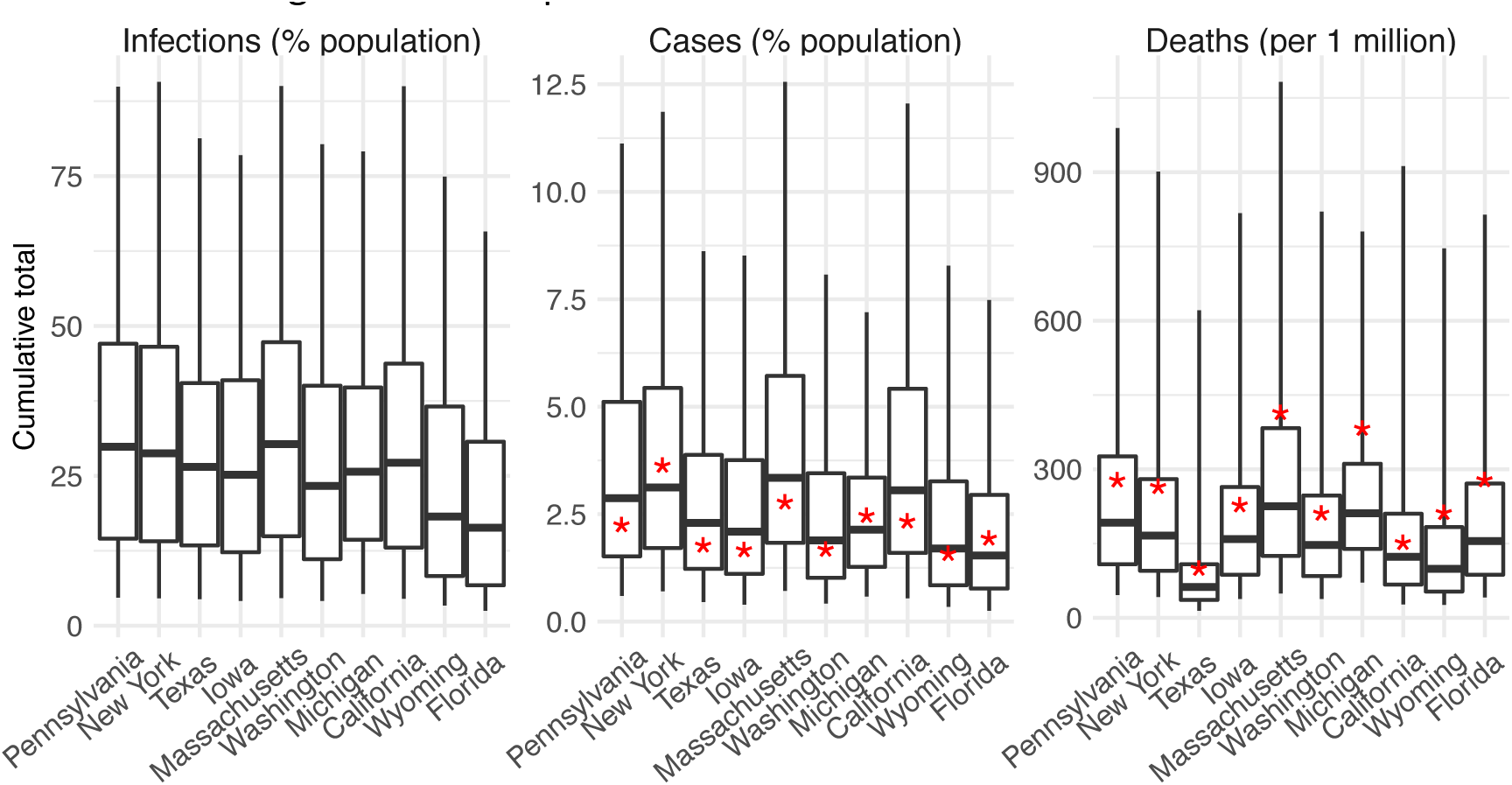
Real-time forecasts of cumulative infections, cases, and deaths during the 2022-2023 respiratory virus season. Box plots show distributions of predicted total number of infections (1^st^ panel, scaled to population size; i.e. attack rate), cases (2^nd^ panel, scaled to population size), and deaths (3^rd^ panel, scaled per 1 million persons) from the week starting 10/2/2022 to the week starting 3/26/2023. Thick line = median; box edge = interquartile range; whisker = 95% prediction interval. The states (x-axis label) are arranged according to accuracy of historical forecast (higher accuracy from left to right). Red asterisks (*) show reported cumulative cases and deaths during the forecast period.

## DISCUSSION

Given the uncertainties surrounding future SARS-CoV-2 transmission dynamics, it is immensely challenging to accurately predict long-lead COVID-19 epidemic outcomes. Here, we have proposed three strategies for sensibly improving long-lead COVID-19 forecast. Retrospective forecast accuracy is substantially improved using the three strategies in combination during both the pre-Omicron and Omicron periods, including for long-lead targets 6 months in the future. This improvement is consistent among 10 representative states across the US, indicating the robustness of the forecast method.

Our first strategy addresses the accumulation of forecast error over time. The simple deflation method proposed here substantially improves forecast accuracy across different model settings (here, different new variant and seasonality forms), time periods (pre-Omicron vs Omicron), and locations (different states). This consistent improvement indicates that deflation is effective in constraining outlier ensemble trajectories. Albeit possibly a severer issue for SARS-CoV-2 due to larger uncertainties and less constrained parameter estimates, error growth is a common challenge in forecasts of infectious diseases not limited to COVID-19 (21, 22). Future work could examine the utility of deflation in improving forecast accuracy for other infectious diseases.

Another challenge facing COVID-19 forecast derives from the uncertainty associated with new variant emergence. Based on past epidemiological dynamics of and population response to SARS-CoV-2 VOCs, we proposed a simple set of heuristic rules and applied them universally across time periods and locations. Despite their simplicity, the results here show that these heuristics substantially improved forecast accuracy compared to forecasts generated without them. These findings suggest that, while it is challenging to forecast the emergence of specific variants, the timing of future variant emergence and the impacts on key epidemiological characteristics (i.e., population susceptibility and virus transmissibility) can be learned from past VOC waves and used to support more accurate forecast. Much uncertainty remains regarding future SARS-CoV-2 genomic evolution and population immunity; however, the heuristics proposed here represent a first step anticipating the dynamic interplay of SARS-CoV-2 new variants and population immunity. Continued work to test the robustness of these heuristics as SARS-CoV-2 and population immunity continue to co-evolve is thus warranted.

The third focus of this study is seasonality. Several prior studies have examined the potential seasonality of COVID-19, using methods such as time series analyses, regression models, and sinusoidal functions (23–25). However, the underlying mechanisms and likely nonlinear response to seasonal drivers are not fully characterized. Several concurrent changes including case ascertainment rate, NPIs and voluntary behavioral changes, and new variants further complicate such characterization. Here, we used local weather data along with a mechanistic model previously developed for influenza (16, 17) to capture the nonlinear response of respiratory virus survival/transmission to humidity and temperature. In addition, we tested an alternative seasonality form given the likely differences between SARS-CoV-2 and influenza (e.g., likely higher infection risk for SARS-CoV-2 than influenza during the summer). When incorporated in a model-inference framework and forecast system, forecasts with both seasonality forms outperformed their counterpart without seasonality. Importantly, improvements during the respiratory season were consistent throughout the pandemic, including for the VOC waves, as well as for all 10 states with diverse climate conditions (Table S4 and Fig S1). These findings indicate the robustness of the seasonality functions and the importance of incorporating seasonality in COVID-19 forecast. More fundamentally, the results support the idea that a common set of seasonal environmental/climate conditions influence the transmission dynamics of respiratory viruses, including but not limited to SARS-CoV-2 and influenza. Among the 10 states tested here, the transformed seasonality function tended to outperform the fixed seasonality function based on parameters estimated for influenza, except for Massachusetts and Michigan (Table S4). This difference in performance suggests there are likely nuances in the seasonality of different respiratory viruses despite shared general characteristics.

To focus on the above three challenges, in our retrospective forecasts, we used data/estimates to account for several other factors shaping COVID-19 dynamics. These included behavioral changes (including those due to NPIs), vaccination uptake, changing detection rates and hence case ascertainment rate, as well as changes in infection fatality risk due to improvement of treatment, vaccination, prior infection, and differences in the innate virulence of circulating variants. For real-time forecast, such data and estimates would likely not be available and thus forecast accuracy would likely be degraded. Nonetheless, as societies emerge from the acute pandemic phase, many of these factors would likely reach certain norms (e.g., a relative stable fraction of the population may continue to adopt preventive measures and detection rates may stay low), reducing these uncertainties. Thus, though demonstrated mainly retrospectively, the superior skill of the forecast methods developed here demonstrate means for generating more accurate and sensible long-lead COVID-19 forecasts.

## METHODS

### Data used for model calibration

We used COVID-19 case and mortality data (26) – adjusted for circulating variants (27, 28) – to capture transmission dynamics, mobility data (29) to represent concurrent NPIs, and vaccination data (30, 31) to account for changes in population susceptibility due to vaccination. For models including seasonality, we used weather data (i.e., temperature and humidity)(32, 33) to estimate the infection seasonality trends. See detailed data sources and processing in the SI.

### Model calibration before forecast generation (i.e. inference)

The model-inference system is similar to systems we developed to estimate changes in transmissibility and immune erosion for SARS-CoV-2 VOCs including Alpha, Beta, Gamma, Delta, and Omicron (17–19). However, to account for the fast waning of vaccine protection against infection and differential vaccine effectiveness (VE) against different variants, here we additionally accounted for variant-specific VE and waning vaccine protection against infection per Eqn 1:

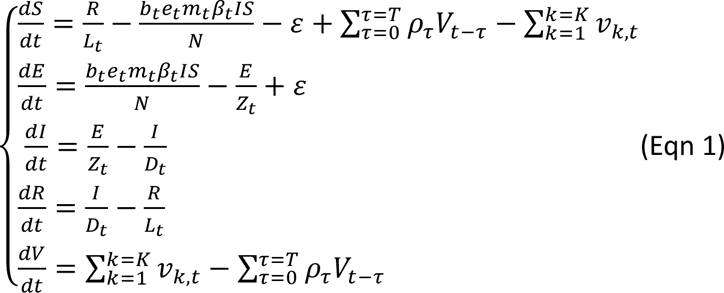

where *S*, *E*, *I*, *R* are the number of susceptible, exposed (but not yet infectious), infectious, and recovered/deceased individuals; *N* is the population size; and *ε* is the number of travel-imported infections. To account for changes due to circulating variants, Eqn 1 includes a time-varying transmission rate β_*t*_, latency period *Z_t_*, infectious period *D_t_*, and immunity period *L_t_*. To account for the impact of NPIs, Eqn 1 uses the relative population mobility (*m_t_*) to adjust the transmission rate and a scaling factor (*e_t_*) to account for potential changes in effectiveness. To account for vaccination and waning, *V* is the number of individuals vaccinated and protected from infection, *v*_*k*,*t*_ is the number of individuals immunized after the *k*-th dose at time *t* and *ρ_τ_* is the probability of losing vaccine protection *τ* days post vaccination (see SI and Table S5). As described below, *b*_*t*_ is the seasonal infection risk at time *t*, depending on the seasonality setting. We further computed the number of cases and deaths each week to match with the observations using the model-simulated number of infections occurring each day (see the SI).

We ran the model jointly with the ensemble adjustment Kalman filter (EAKF (34)) and weekly COVID-19 case and mortality data to estimate the model state variables (e.g., *S*, *E*, and *I*) and parameters (e.g., β_*t*_, *Z_t_*, *D_t_*, *L_t_*, *e_t_*). Briefly, the EAKF uses an ensemble of model realizations (*n*=500 here), each with initial parameters and variables randomly drawn from a *prior* range (Table S5). After model initialization, the system integrates the model ensemble forward in time for a week (per Eqn 1) to compute the prior distribution for each model state variable, as well as the model-simulated number of cases and deaths for that week. The system then combines the prior estimates with the observed case and death data for the same week to compute the posterior per Bayes’ theorem (34). In addition, as in (17–19), during the filtering process, we applied space-reprobing (35), i.e., random replacement of parameter values for a small fraction of the model ensemble, to explore a wider range of parameter possibilities (Table S5). The space-reprobing algorithm, along with the EAKF, allows the system to capture potential changes over time (e.g., increased detection for variants causing more severe disease, or increases in population susceptibility and transmission rate due to a new variant).

### Variations in forecast systems (deflation, new variant, and seasonality settings)

In total, 12 forecast approaches were tested (3 deflation levels × 2 new variants settings × 3 seasonality forms). The deflation algorithm is patterned after covariance inflation, as used in filtering methods (9, 10). However, unlike inflation applied during filtering (i.e., the model training period via data assimilation), deflation is applied during the forecast period. As the state variables change dynamically per the epidemic model (i.e., Eqn 1), the error of some epidemic trajectories can amplify exponentially over time. Thus, here we applied deflation only to the state variables (i.e., not to the model parameters), per:

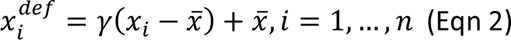

where *x_i_* is *i*-th ensemble member of a given state variable (here, S, E, or I) at each time step during the forecast period, before the deflation; 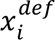 is the corresponding “deflated” value; γ is the deflation factor; and 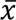 is the ensemble mean of the state variable. Per Eqn 2, deflation retains the ensemble mean, while reducing the ensemble spread to constrain error accumulation. In this study, we tested three levels of deflation, by setting γ to 1 (i.e., no deflation), 0.95, and 0.9, separately.

To anticipate and account for potential surges and their impact on COVID-19 epidemic outcomes, we tested two approaches. The first, baseline approach simply assumes there are no changes in the circulating variant during the forecast period. For this approach, the forecasts were generated using the latest population susceptibility and transmission parameter estimates at the point of forecast initiation. For the second, new variant approach, we devised a set of heuristics to anticipate the likely timing and impact of new variant emergence during the forecast period, as detailed in the SI.

Seasonality is incorporated in the epidemic model (Eqn 1) and applied throughout the model calibration (i.e., inference) and forecast periods. Here, we tested three seasonality settings. The first assumes no changes in seasonal risk of infection, by setting *b_t_* in Eqn 1 to 1 for all weeks (referred to as “no seasonality”). The second seasonality form (termed “fixed seasonality) estimates the relative seasonality trend (*b_t_*, same as in Eqn 1) using local humidity and temperature data, based on the dependency of respiratory virus survival, including that of SARS-CoV-2, to temperature and humidity (12, 17, 18, 36); see Eqn 3 and details in the SI. The third seasonality setting (termed “transformed seasonality”) transforms the *b_t_* estimates to allow flexibility in the seasonal trend, including the peak timing, the number of weeks during a year with elevated infection risk, and the lowest risk level (see Eqn 4 and details in the SI). Due to the lack of SARS-CoV-2 data to inform the parameter estimates, here we opted to optimize the range for each parameter and used the best parameter ranges (see the SI and Fig S5) in the transformed seasonality model in the main analysis.

### Retrospective forecast

We tested the above 12 model-inference and forecast approaches (3 deflation levels × 2 new variants settings × 3 seasonality forms) for 10 states, i.e., California, Florida, Iowa, Massachusetts, Michigan, New York, Pennsylvania, Texas, Washington, and Wyoming. The 10 states span the 10 Health and Human Services (HHS) regions across the US, representing a wide range of population characteristics and COVID-19 pandemic dynamics (Fig 1). For all states, we generated retrospective forecasts of weekly cases and deaths 26 weeks (i.e., 6 months) into the future for the non-Omicron period and the Omicron period, separately. For the non-Omicron period, we initiated forecasts each week from the week of July 5, 2020 (i.e., after the initial wave) through the week of August 15, 2021. Note that because each forecast spans 6 months, the last forecasts initiated in mid-August 2021 extend to mid-Feb 2022, covering the entire Delta wave (see Supplemental text for details). For the Omicron period, we initiated forecasts starting 5 weeks after local detection of Omicron BA.1 (roughly in early December 2021, depending on local data) through the week of September 25, 2022 (i.e., the last week of this study).

To generate a forecast, we ran the model-inference system until the week of forecast, halted the inference, and used the population susceptibility and transmissibility of the circulating variant estimated at that time to predict cases and deaths for the following 26 weeks (i.e., 6 months). Because the infection detection rate and infection-fatality risk are linked to observations of cases and deaths (see the SI), changes of these quantities during the forecast period could obscure the underlying infection rate and forecast accuracy. Thus, for these two parameters specifically, we used available model-inference estimates for corresponding forecast period weeks to allow comparison of model-forecast cases and deaths with the data while focusing on testing the accuracy of different model settings (e.g., seasonality and new variant settings). For the same reason, we used all available mobility and vaccination data including those for the forecast period, which would not be available in real time. For weeks in the future without data/estimates, we used the latest estimates instead. To account for model and filter stochasticity, we repeated each forecast 10 times, each time with initial parameters and state variables randomly drawn from the same prior ranges.

To evaluate forecast performance, we computed both the log score based on the probabilistic forecast and the accuracy of point prediction for 1) 1- to 26-week ahead prediction, and 2) peak week, 3) peak intensity, and 4) cumulative total over the 26-week forecast period, for cases and deaths, separately. Details are provided in the SI. Here, in brief, to compute the log score, we first binned the forecast ensemble to generate the forecast probability distribution Pr(*x*), and took the logarithm of the sum of Pr(*x*) across all related bins including the one including the observation (*bin*^∗^) and two adjacent ones (*bin*^∗–1^ and *bin*^∗+1^):

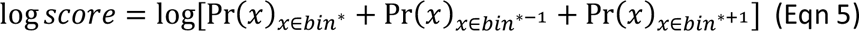

For the accuracy of point prediction, we deemed a forecast accurate (assigned a value of 1) if the median of the forecast ensemble is within ±1 week of the observed peak week or within ±25% of the observed case/death count, and inaccurate (assigned a value of 0) otherwise. As noted above, when aggregated over multiple forecasts, the average would represent the percentage of time a point prediction is accurate within these tolerances.

We compared the performance of each forecast approach (i.e., each of the 12 combinations of deflation, new variant setting, and seasonality form) overall or by forecast target, segregated by time period or respiratory virus season. To compute the overall score for each stratum, we took the arithmetic mean of the log score or point prediction accuracy of forecasts generated by each forecast system, either across all forecast targets or for each target, over 1) all forecast weeks during the entire study period (i.e., July 2020 – September 2022), 2) the pre-Omicron period and Omicron period, separately, 3) the respiratory virus season (mid-October to mid-April, 6 months) and off season (the remaining 6 months), separately.

For pairwise comparison of forecast approaches, we computed the difference of log score or accuracy by simple subtraction of the two arithmetic-means. Relative difference was also computed. For the log score, the percent relative difference was computed as:

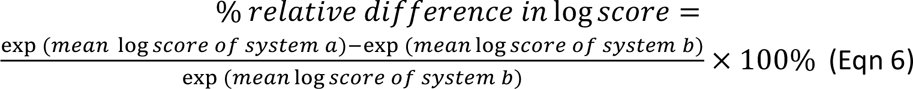

As noted in (37), the exponent of the mean log score (Eqn 5) can be interpreted as the probability correctly assigned to the bins containing the observations; thus Eqn 6 gives the relative difference in the correctly assigned forecast probability. The percent relative difference in accuracy of point prediction was computed as:

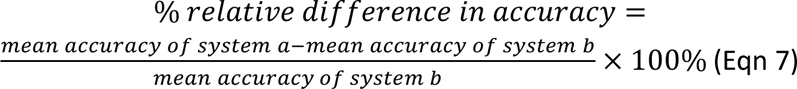

In addition, we also computed the pair-wised difference of log score or accuracy (paired by forecast week for each target and location) and used boxplots to examine the distributions (see, e.g., Fig 6). We used the Wilcoxon rank sum test, a non-parametric statistical method, to test whether there is a difference in the median of the pair-wised differences (38).

### ARIMAX model forecast for comparison with approaches developed in this study

Auto-Regressive Integrated Moving Average (ARIMA) models and ARIMAX models (X represents external predictors) are commonly used to forecast different outcomes. For comparison with the approaches developed here, we also tested five ARIMA(X) models and used them to generate retrospective forecasts per the same procedure described above. The first model (i.e., simple ARIMA model) used weekly case or mortality data alone for model training. The remaining four models were ARIMAX models: 1) using case/mortality data and mobility data including for the forecast period (i.e., X = mobility; referred to as “ARIMAX.MOB”); 2) using case/mortality data and the estimated seasonal trend from the fixed seasonal model (i.e., X = seasonality; referred to as “ARIMAX.SN”); 3) using case/mortality data, mobility data, and the estimated seasonal trend (i.e., X = mobility and seasonality; referred to as “ARIMAX.MS”); and 4) using case/mortality data, mobility data, the estimated seasonal trend, and vaccination data (i.e., X = mobility, seasonality, and vaccination; referred to as “ARIMAX.FULL”). For vaccination, to account for the impact accumulated over time, we used cumulative vaccinations (here, in the past 3 months for cases, and the past 9 months for deaths). For model optimization, we used the “auto.arima” function of the “forecast” R package (39), which searches all possible models within the specified order constraints (here, we used the default settings) to identify the best ARIMA(X) model (here, based on the corrected Akaike information criterion by default).

For the five model forms, the ARIMAX.FULL model (i.e., including mobility, seasonality, and vaccination) was only able to generate forecasts for less than half of the study weeks as the auto.arima function was unable to identify parameters for this model. The other four models were able to generate forecasts for most study weeks and across the entire study period, the ARIMAX.SN (i.e., seasonality included) performed the best (see Table S5). As such, we used the ARIMX.SN model as a benchmark model for comparison with approaches developed in this study.

### Preliminary assessment of real-time forecasts for the 2022 – 2023 respiratory virus season

The last forecasts in this study were generated using all available data up to the week starting October 2, 2022 and spanned 6 months through the week starting March 26, 2023. These were real-time forecasts generated without future information. We assess these real-time forecasts using data downloaded on March 31, 2023 (1 day after the data release). Since these data may be revised in the future (*n.b.* data revision after the initial release has been common), we consider the assessment preliminary. As detailed in the SI, we used case and mortality data from the New York Times (NYT; (26)) for model calibration prior to generating the forecasts. However, in the six months since the initial study, NYT data have become more irregular for some states, likely due to infrequent data reporting and updating. As such, for this preliminary assessment, we instead used data from the Centers for Disease Control and Prevention (CDC)(40), except for mortality in Washington State for which the CDC data appeared to be misdated whereas NYT data and mortality data from the Center for Systems Science and Engineering (CSSE) at Johns Hopkins University (41) were consistent with each other. In addition, the CDC data were aggregated for each week from Thursday to Wednesday, rather than Sunday to Saturday. To enable the comparison, we thus shifted the dates of the CDC data 3 days.

All inference, forecast, and statistical analyses were carried out using the R language (https://www.r-project.org).

## Data Availability

Data and model code are publicly available at https://github.com/wan-yang/covid_long_lead_forecast

## Acknowledgements

This study was supported by the National Institute of Allergy and Infectious Diseases (AI145883 and AI163023), the Centers for Disease Control and Prevention (CDC) and the Council of State and Territorial Epidemiologists (CSTE; contract no.: NU38OT00297), and the CDC Center for Forecasting and Outbreak Analytics (contract no.: 75D30122C14289).

## Supporting Information (SI)

### Supplemental methods

Addition details on 1) Data sources and processing; 2) Modeling of variant-specific vaccine effectiveness and waning vaccine protection against infection; 3) Observation model to account for under-detection and time-lags in COVID-19 outcomes; 4) Settings for anticipating the impact of new variants (the new variant approach); 5) The fixed seasonality model; 6) The transformed seasonality model; and 7) The retrospective forecast and forecast evaluation.

## SUPPLEMENTAL METHODS

### Data sources and processing

For model calibration, we used reported COVID-19 case and mortality data to capture transmission dynamics, mobility data to represent concurrent NPIs, and vaccination data to account for changes in population susceptibility due to vaccination. State level COVID-19 case and mortality data were sourced from the New York Times (NYT) (1) and included all variants. In our previous studies, overall case/mortality data were sufficient to estimate key epidemiological parameters when different VOC waves were separated in time (2–4); however, in the US, the Omicron BA.1 wave overlapped substantially with the Delta wave during November 2021 – January 2022, making inference challenging. Thus, here we separated the forecasts into two periods (i.e., a pre-Omicron period combining all non-Omicron variants, and an Omicron period combining all Omicron subvariants) and used variant-specific case and mortality data for model training and forecast evaluation for each period. Specifically, we used variant proportion data sourced from GISAID (5) and compiled by CoVariants.org (6) to compute the weekly number of cases and deaths due to non-Omicron variants and Omicron, separately (for simplicity, loosely referred to as variant-specific case and mortality data). Because only biweekly variant proportion data at the state level were available from CoVariants.org, we used a spline function to impute weekly variant proportion. To compute weekly variant-specific cases, we multiplied the NYT weekly case data by the estimated weekly variant proportion for the same week. To compute weekly variant-specific deaths, we multiplied the NYT weekly mortality data by the estimated weekly variant proportion three weeks later (i.e., assuming a 3-week lag from case detection to death; note the 3-week lag was based on the approximate time lag between the peaks of incidence and mortality time series).

Mobility data were derived from Google Community Mobility Reports (7); we aggregated all business-related categories (i.e., retail and recreational, grocery and pharmacy, transit stations, and workplaces) in all locations in each state to weekly intervals. State level COVID-19 vaccination data were sourced from Our World in Data (8, 9). For models including seasonality, weather data (i.e., temperature and humidity) were used to estimate infection seasonality trends. Hourly surface station temperature and relative humidity came from the Integrated Surface Dataset (ISD) maintained by the National Oceanic and Atmospheric Administration (NOAA) and are accessible using the “stationaRy” R package (10, 11). We computed specific humidity using temperature and relative humidity per the Clausius-Clapeyron equation (12). We then aggregated these data for all weather stations in each state with measurements since 2000 and calculated the average for each week of the year during 2000-2020.

### Modeling variant-specific vaccine effectiveness (VE) and waning vaccine protection against infection

As noted in the main text, the epidemic model in Eqn 1 includes vaccination and waning vaccine protection. Specifically, vaccination including boosters is represented using the term 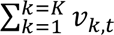, where *v*_k,*t*_ is the number of individuals immunized at time *t*, after the *k*-th dose (*k* = 1,…,3 for 2 primary and 1 booster dose here, excluding those immunized after previous doses). We computed *v*_k,*t*_ using vaccination data and adjusted for the delay in antibody development (here, 14 days for the 1^st^ dose and 7 days for subsequent doses) and variant specific VE (13–17). Note that while the 2^nd^ booster dose has been administered for a subset of the population, such data have not been made publicly available and thus not included in our model. Further, given the 2^nd^ dose and 3^rd^ dose (i.e. 1^st^ booster) were administered ∼6 months apart (i.e., beyond the estimated VE duration against infection (16)), here we combined data for these two doses. In doing so, we have simplified the model and implicitly assumed that the 3^rd^ dose resumed VE against infection to a level similar to the 2^nd^ dose, as the same VE was applied (Table S8).

The model further accounts for waning of vaccine protection against infection, using the term 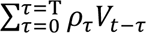. We computed the total number who were vaccinated *τ* days ago and lost protection on day-*t* (*V_t-τ_*) per the VE waning probability (*ρ_τ_*). The probabilities *ρ_τ_* for time *τ* =0, …, T (T= the maximum duration from the earliest vaccination rollout; Table S8) were calculated using VE duration data (16) and *V_t-τ_* was computed per line 5 of Eqn 1.

### Observation model to account for under-detection and time-lags in COVID-19 outcomes

We computed the number of cases and deaths each week using the model-simulated number of infections occurring each day to match with the observations, as done in Yang et al. (18). Briefly, we included 1) a time-lag from infectiousness to detection (i.e., an infection being diagnosed as a case), drawn from a gamma distribution with a mean of *T_d,mean_* days and a standard deviation of *T_d, sd_* days, to account for delays in detection; 2) an infection-detection rate (*r_t_*), i.e. the fraction of infections (including subclinical or asymptomatic infections) reported as cases, to account for under-detection; 3) a time-lag from infectiousness to death; and 4) an infection-fatality risk (*IFR_t_*). Each week, the infection-detection rate (*r_t_*), infection-fatality risk (*IFR_t_*), and the two time-to-detection parameters (*T_d, mean_* and *T_d, sd_*) were estimated along with other parameters (see main text). The time-lag from infectiousness to death during the pre-Omicron period was drawn from a gamma distribution with a mean of 14 days and a standard deviation of 14 days, roughly based on data from New York City (unpublished work). For the Omicron period, many deaths were identified posthumously; thus, it is difficult to estimate the time-lag from infection to death for Omicron infections. Here, based on the slightly longer time-lag between the peaks of case and mortality time series during the Omicron period, we assumed a gamma distribution with a mean of 24 days (i.e., assuming an additional 10-day lag) and a standard deviation of 14 days.

To compute the model-simulated number of new cases each week, we multiplied the model-simulated number of new infections per day by the infection-detection rate, and further distributed these simulated cases in time per the distribution of time-from-infectiousness-to-detection. Similarly, to compute the model-simulated deaths per week and account for delays in time to death, we multiplied the simulated-infections by the IFR and then distributed these simulated deaths in time per the distribution of time-from-infectious-to-death. We then aggregated these daily numbers to weekly totals to match with the weekly case and mortality data for model inference, as described in the main text.

### Settings for anticipating the impact of new variants (the new variant approach)

The uncertainty due to the possible emergence or surge of new variants in the future is a major challenge for long-lead COVID-19 forecast. To address this challenge, we devised a set of heuristics to anticipate the likely timing and impact of new variant emergence during the forecast period (i.e., the new variant approach). For the very near future (1- to 5 weeks), we used available genomic sequencing data (see “Data sources and processing”). Specifically, we first estimated the growth rate for each circulating variant based on variant proportion 6- to 2 weeks prior to the week of forecast initiation (i.e., assuming a 2-week lag for genomic data collection); for simplicity, we used a log-linear model [i.e., log(variant proportion during week-*t*) ∼ week-*t*]. If any variant had a high growth rate (here, arbitrarily set to 10% per week), we deemed it a rising variant that could further affect the population susceptibility and overall virus transmissibility. To anticipate its impact, we then used a smoothing spline to 1) project when the variant would reach a 100% proportion and 2) project the number of weeks for the variant to grow from 0% to 100%. The first estimate was then used to set the timing of the continued impact and the second estimate was used to scale the increases in population susceptibility. Here, arbitrarily, we assumed a baseline of 1.5 – 4.5% increase in susceptibility for each week the new variant increased in proportion; however, if the estimated growth rate (*g*) was >20%, to account for the faster growth, we scaled that baseline by a factor of (1+*g*)/1.2. While the growth advantage of a new variant could also come from increased transmissibility, for simplicity, here we opted to solely adjust for population susceptibility. In addition, given the fast displacement of new variants, we opted not to use projected estimates 5 weeks beyond the forecast week, i.e. these changes were only applied to the first 5 weeks of a forecast.

When genomic data could not be used (i.e. beyond the first 5 forecast weeks or when genomic data were not available), we used the following heuristics to anticipate the likely timing and impact of a new variant surge:

1. New variants tend to emerge after a recent large wave (here, defined arbitrarily as a 25% attack rate over 3 months for the non-Omicron period, and a 33% attack rate over 2 months for the Omicron period). We identified these times using estimated/forecasted infection rates during the preceding months and the forecast period.
2. New variants tend to emerge and/or become widespread during northern hemisphere winter (December – February), southern hemisphere winter (June – August), and/or the monsoon season (e.g. June – September in India) and could be introduced to the US during these months. We identified these times using calendar month.
3. During the above times with potential new variant emergence, the population susceptibility and virus transmissibility could increase. Accordingly, to account for susceptibility changes, we resampled half of the model ensemble to increase the population susceptibility by 2-9% for weeks flagged per the conditions described in i and ii. However, this susceptibility increase was only triggered when the mean population susceptibility was below 40% to avoid over-adjustment. Similarly, to avoid the system being trapped in an outbreak-begets-outbreak cycle, no further adjustments were made to susceptibility if a wave had been forecast the prior weeks or the cumulative adjustment had exceeded a threshold (here, set to 40% of the population over 26 weeks for pre-Omicron period and 60% for the Omicron period). To account for transmissibility changes, we expanded the variance of the transmission rate (i.e., β_*t*_ in Eqn 1) by applying an inflation factor of 1.3 (pre-Omicron period) or 1.1 (Omicron period) to ensemble members falling between the 50^th^ and 95^th^/90^th^ (pre-Omicron/Omicron period) percentiles (i.e., the ones with higher but not too extreme values) for weeks identified per the conditions described in i and ii.

### The fixed seasonality model

The fixed seasonality model represents the dependency of respiratory virus survival, including that of SARS-CoV-2, to temperature and humidity (19, 20) per the following equations:

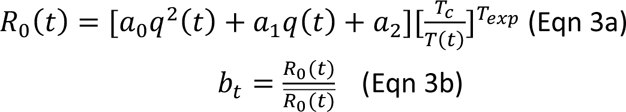

As described previously (3, 4), the seasonality function in Eqn 3a assumes that humidity has a bimodal effect on seasonal risk of infection, with both low and high humidity conditions favoring transmission [i.e., the parabola in the 1st set of brackets, where *q*(*t*) is weekly specific humidity measured by local weather stations and *t* = 1,…,52, i.e., week 1 to week 52 of the year]; this effect is further modulated by temperature, with low temperatures promoting transmission and temperatures above a certain threshold limiting transmission [i.e., the 2nd set of brackets, where *T*(*t*) is weekly temperature measured by local weather stations and *T_c_* is the threshold]. As SARS-CoV-2 specific parameters (*a*_0_, *a*_1_, *a*_2_, *T_c_*, and *T_exp_* in Eqn 3a) are not available, we used parameters estimated for influenza (21) and scaled the weekly outputs [i.e., *R*_0_(*t*)] by the annual mean (i.e., 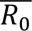) per Eqn 3b, as done in Yang and Shaman (4). In doing so, the scaled outputs (*b_t_*) are no longer specific to influenza; rather, they represent the *relative*, seasonality-related transmissibility by week, general to viruses sharing similar seasonal responses. The estimated relative seasonal trend, *b_t_*, is then used to adjust the relative transmission rate at time *t* in Eqn 1.

### The transformed seasonality model

The transformed seasonality model transforms the *b_t_* estimates from Eqn 3b to allow flexibility in the seasonal trend. To do so, we include three parameters to fine tune the peak of the seasonal trend (*p_shift_*; i.e., the number of weeks earlier or later than the peak estimated for influenza), the number of weeks during a year with *b_t_* >1 (δ; i.e., the duration with elevated infection risk), and another parameter *b*_*t,lwr*_ that adjusts the lowest *b_t_* value. Specifically, the transformation first adjusts values of *b_t_* greater than 1, by shifting the timing by *p_shift_* weeks and adjusting the duration with elevated infection risk to δ, per

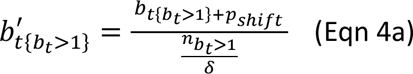

where *n*_*b*_*_t_*>1 is the number of weeks with *b_t_* >1 during the 1-year cycle. For weeks with *b_t_* ≤1, the transformation adjusts the values, by shifting the timing by *p*_*shift*_ weeks and adjusting the duration with lower infection risk to 52 − δ, per

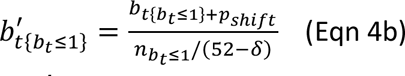

The approach then further scales 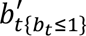 to increase the relative infection risk, per

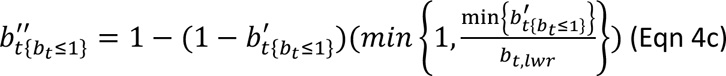

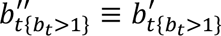 and 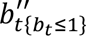 are then pooled together and scaled to have a mean of 1 over

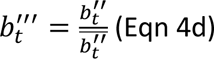

There could be multiple combinations of the three parameters (i.e., *p_shift_*, δ, and *b*_*t,lwr*_). Due to the lack of SARS-CoV-2 data to inform the parameter estimates, here we opted to optimize the range for each parameter (as opposed to estimate specific best-fit parameters). Briefly, we tested 2 levels (low vs. high) for each parameter and thus 8 in combination for each state (Fig S5). We then identified the best range for each state based on forecast performance during the 2^nd^ wave, i.e., before the surge of SARS-CoV-2 VOCs to minimize potential confounding. The best parameter ranges (Fig S5) were then used in the transformed seasonality model in the main analysis.

### Additional details on the retrospective forecast and forecast evaluation

As noted in the main text, retrospective forecasts for the non-Omicron period were done through the week of August 15, 2021. We stopped initiating the non-Omicron forecasts in mid-August 2021 to allow at least a few weeks of Delta-related data to calibrate the model before forecasting the Delta wave. However, a 6-month forecast initiated during mid-June – mid-August 2021 would extend to mid-December 2021 – mid-Feb 2022, when Omicron BA.1 had become predominant, depending on location; this overlap would lead to lower forecast accuracy, since here we did not account for the emergence of Omicron BA.1 and fast displacement of Delta. Given the low number of Delta-associated cases/deaths in 2022, weekly targets (i.e., 1- to 26-week ahead prediction) for weeks in 2022 were excluded from the evaluation; however, as Delta was the main circulating variant during the 6-month period for these forecasts, all the overall targets (i.e., peak week, peak intensity, and cumulative total) were evaluated based on Delta-specific data and included in the analysis.

Both the model inference and forecast were run with *n* = 500 model realizations (i.e., ensemble members). The ensemble and its distribution provided probabilistic forecasts for 4 types of targets here, i.e., 1-to 26-week ahead prediction, peak intensity, peak week, and cumulative totals over the entire 26-week forecast period. For example, for the 1-week ahead prediction (*c*_*t*+1_), the fraction of ensemble members falling in a given bin [*c_i_*, *c_i_*_+1_) can be used to represent the forecast probability density, i.e., 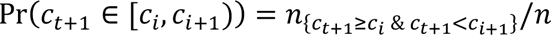 Similarly, for the peak week prediction, predicted peak week by individual ensemble members (*p_w_* = 1, 2, …, 26) can be aggregated and the distribution can be used to represent the probability distribution of the forecast, i.e., 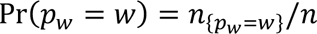.

The forecast probabilities can then be used to compute the log score for evaluation. To do so, we first binned the forecast ensemble to generate the forecast probability distribution Pr(*x*), e.g., Pr(*c*_*t*+1_) for 1-week ahead prediction and Pr(*p_w_*) for peak week. Here, for cases, bins of the weekly targets were set to [0, 0.05%), [0.05%, 0.1%), …, [0.95, 1%), and [1%, 100%] (i.e., increments of 0.05%, or 500 per million people, up to 1% of the population; and the rest combined in the last bin); bins of cumulative cases over 26 weeks were set to [0, 2%), [2%, 4%), …, [8%, 10%), [10%, 15%), [15%, 20%),…,[45%, 50%), and [50%, 100%] (i.e., increments of 2% up to 10%, then increments of 5% up to 50% of the population; and the rest combined in the last bin). For mortality, bins of the weekly targets were set to [0, 0.001%), [0.001, 0.002), …, [0.019%, 0.02%), and [0.02%, 100%] (i.e., increments of 0.001%, or 10 per million people, up to 0.02% of the population; and the rest combined in the last bin); bins of cumulative deaths over 26 weeks were set to [0, 0.02%), [0.02%, 0.04%), …,[0.08%, 0.1%), [0.1%, 0.15%), [0.15%, 0.2%),…, [0.45%, 0.5%), and [0.5%, 100%] (i.e., increments of 0.02% up to 0.1%, then increments of 0.05% up to 0.5% of the population; and the rest combined in the last bin). For the peak week of both cases and deaths, the bin size was set to 1 week. The log score was then computed as:

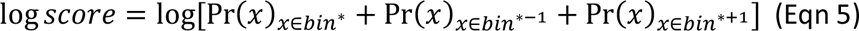

where Pr(*x*) is the forecast probability for target *x*; *bin*^∗^ is the bin that contains the observed value for that target (see bin specifications above) and *bin*^∗-1^ and *bin*^∗+1^ are the two adjacent bins. Note that, here we used smaller bins and deemed ensemble members falling within the bin covering the observation and its two adjacent bins accurate, which is equivalent to using a single larger bin spanning all those smaller bins. However, as the probabilistic forecasts (i.e., probabilities in each bin) were generated and stored before the final evaluation, using smaller bins allowed more flexible post processing and evaluation if needed (e.g., the log score can be computed based on a single small bin if preferred).

---

**Fig S1.**
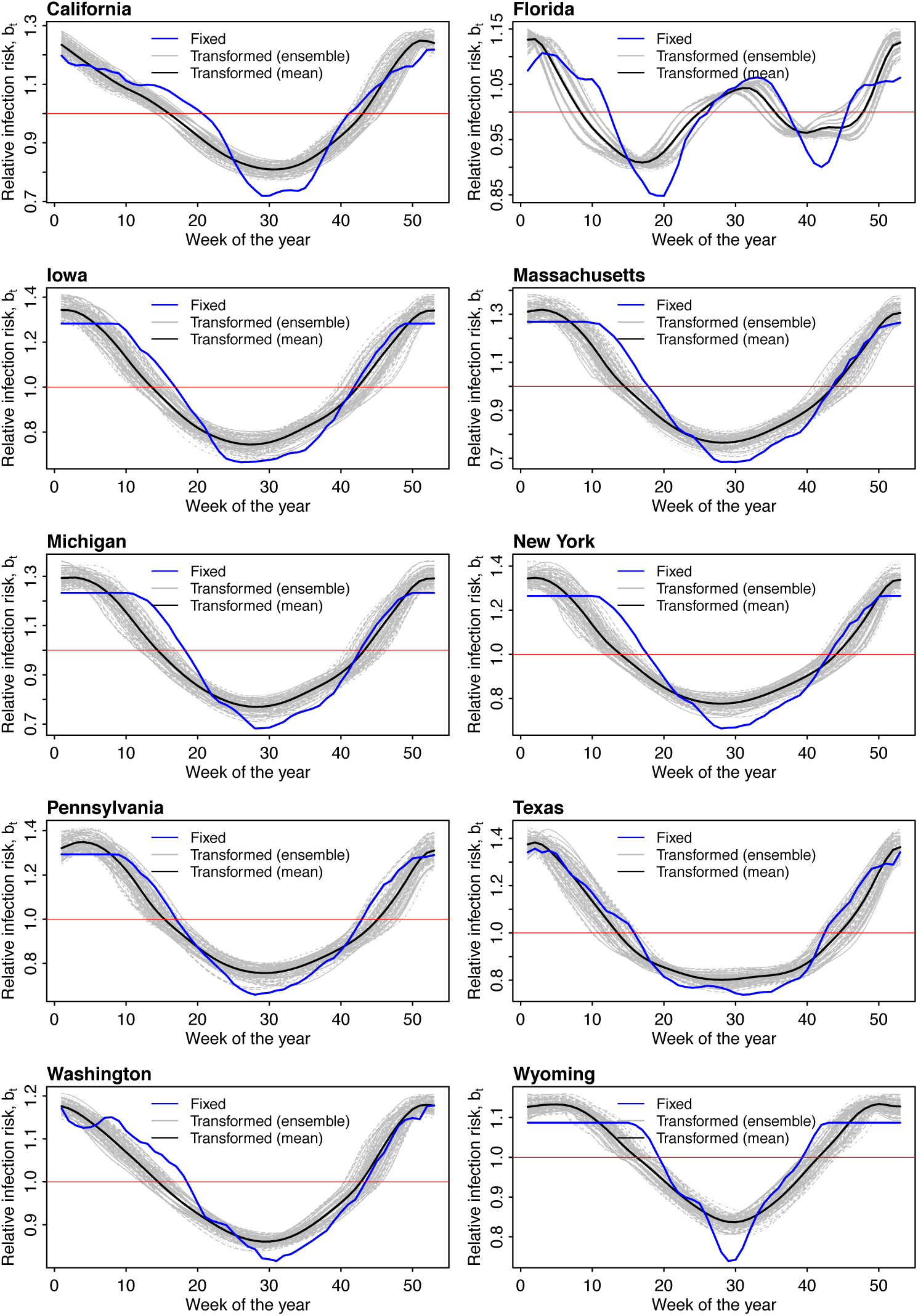
Comparison of seasonality forms. For each state (each panel), the blue line shows the estimated trend of seasonal infection risk using Eqns 3a-b and location weather data (temperature and humidity). Grey lines show 100 examples of the transformed seasonal trends per Eqns 4a-d with parameters randomly sampled from the best parameter ranges (Fig S4); the black line shows the mean of the 100 example trends.

**Fig S2.**
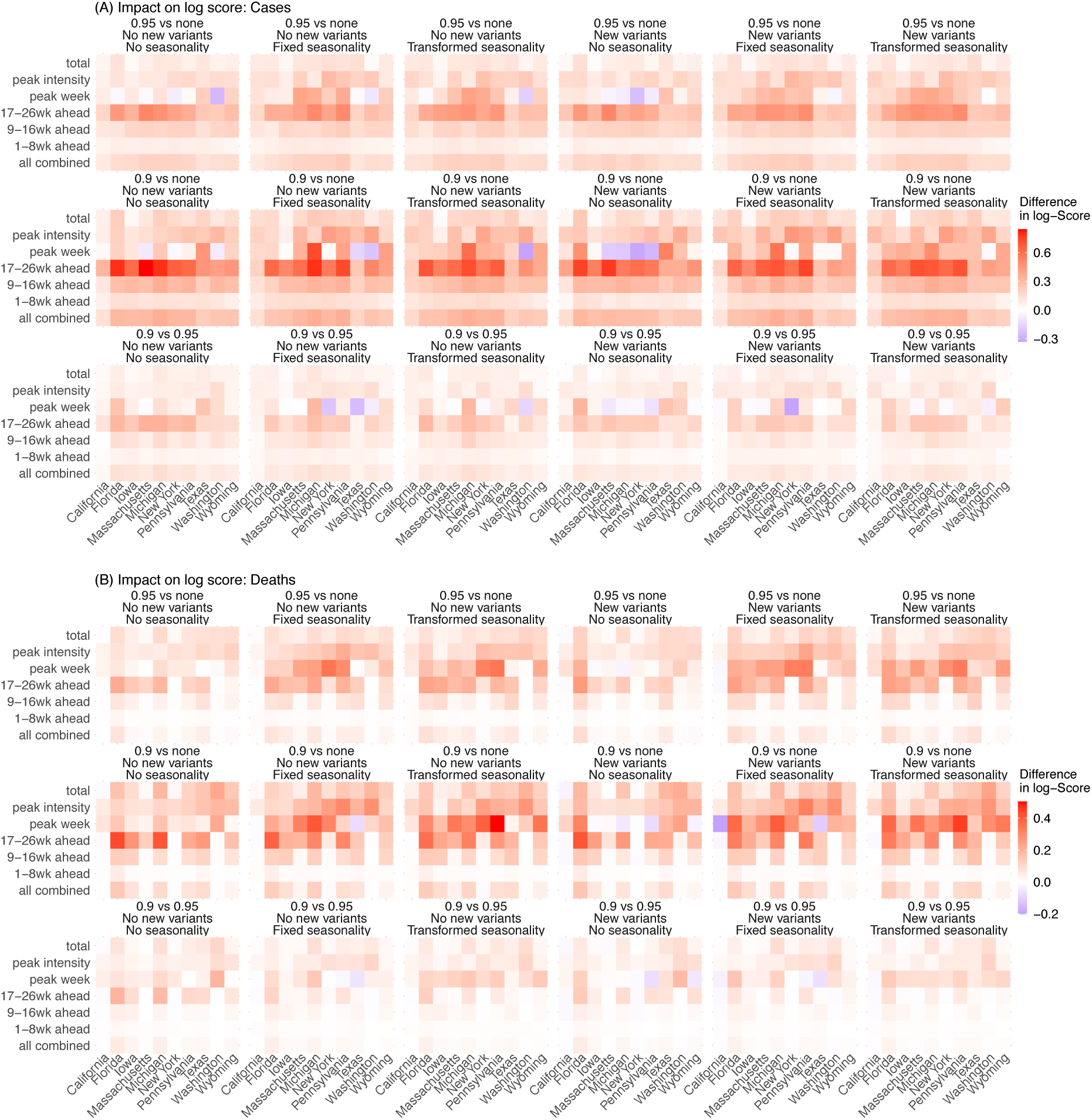
Impact of deflation on probabilistic forecast of different targets. Heatmaps show differences in mean log score for cases (A) and deaths (B), between each forecast approach with different deflation settings (deflation factor γ = 0.95 vs none in the 1^st^ row, 0.9 vs none in the 2^nd^ row, and 0.9 vs 0.95 in the 3^rd^ row; see panel subtitles). Results are aggregated over all forecast weeks for each type of target (y-axis), forecast approach (see specific settings of new variants and seasonality in subtitles), and location (x-axis). For each pairwise comparison (e.g., 0.95 vs none), a positive difference indicates the former approach (e.g., 0.95) outperforms the latter (e.g., none).

**Fig S3.**
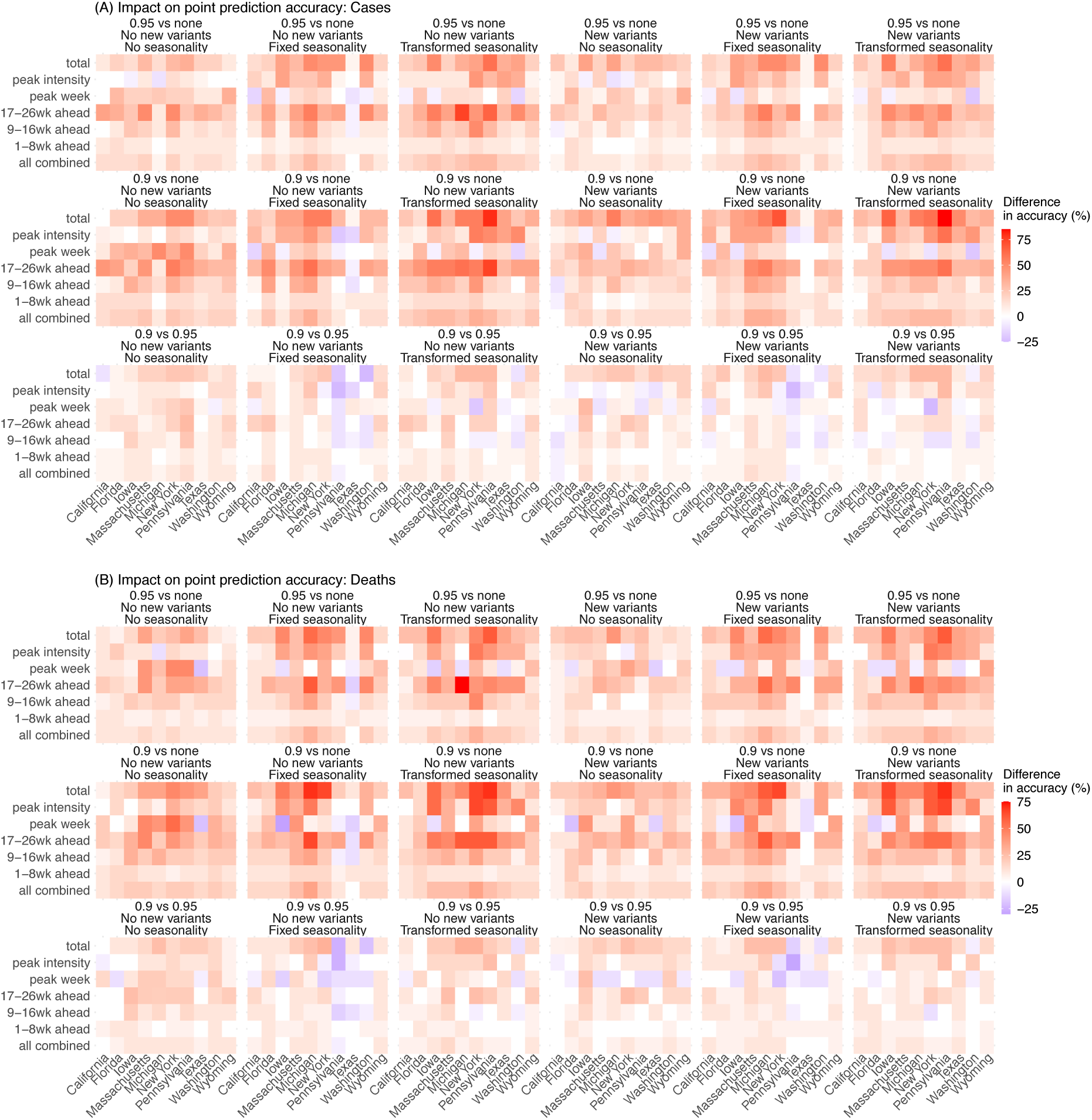
Impact of deflation on point estimate accuracy of different targets. Heatmaps show differences in forecast accuracy of point estimates for cases (A) and deaths (B), between each forecast approach with different deflation settings (deflation factor γ = 0.95 vs none in the 1^st^ row, 0.9 vs none in the 2^nd^ row, and 0.9 vs 0.95 in the 3^rd^ row; see panel subtitles). Results are aggregated over all forecast weeks for each type of target (y-axis), forecast approach (see specific settings of new variants and seasonality in subtitles), and location (x-axis). For each pairwise comparison (e.g., 0.95 vs none), a positive difference indicates the former approach (e.g., 0.95) outperforms the latter (e.g., none).

**Fig S4.**
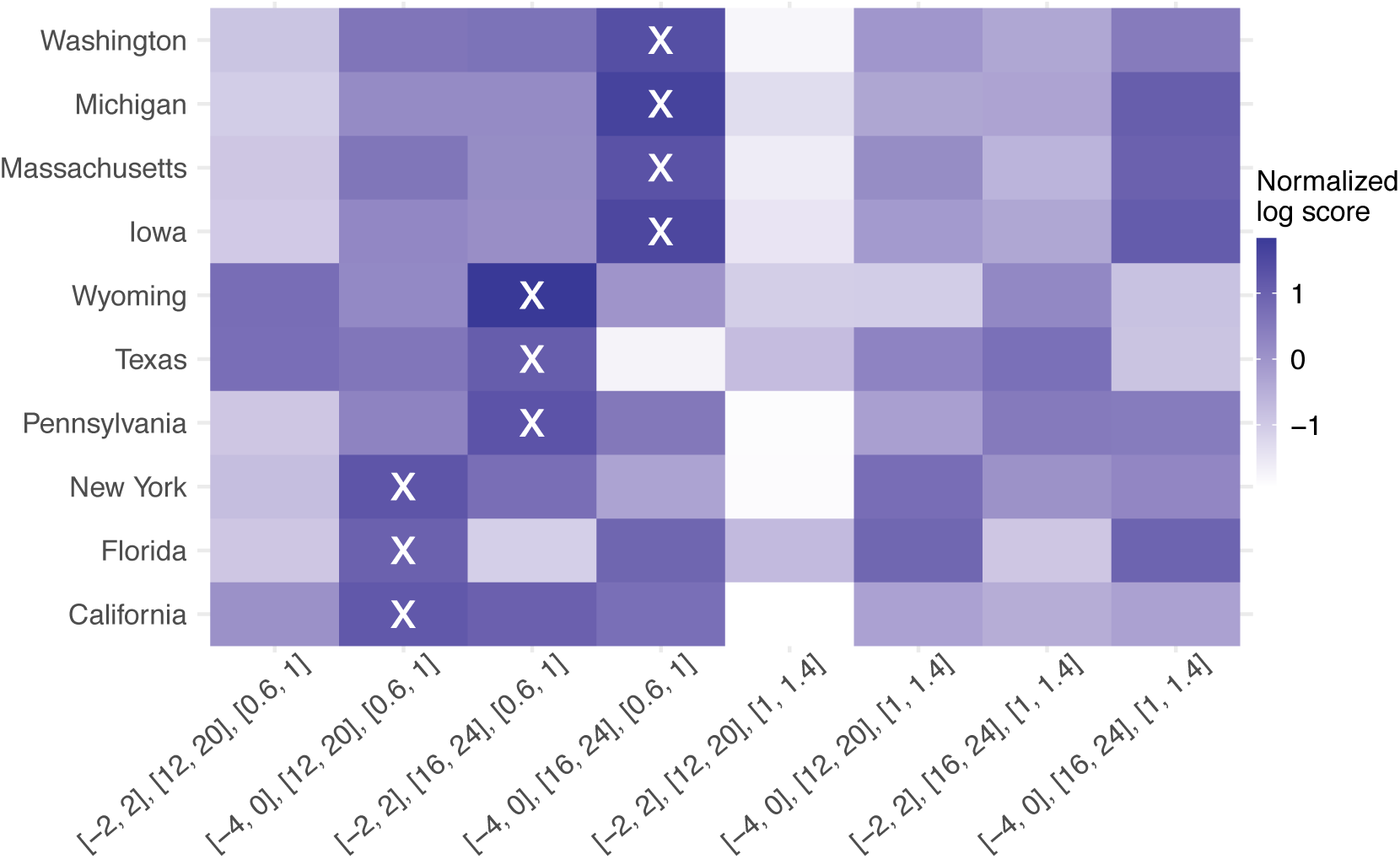
Comparison of forecast performance using the transformed seasonality function, with different parameter ranges. The parameter ranges are shown in x-axis labels for the three parameters in Eqn4a-d (from bottom to top: *p_shift_*, *δ*, and *b_t, lwr_*). ‘x’s indicate the best parameter ranges for the corresponding state.

**Table S1.**
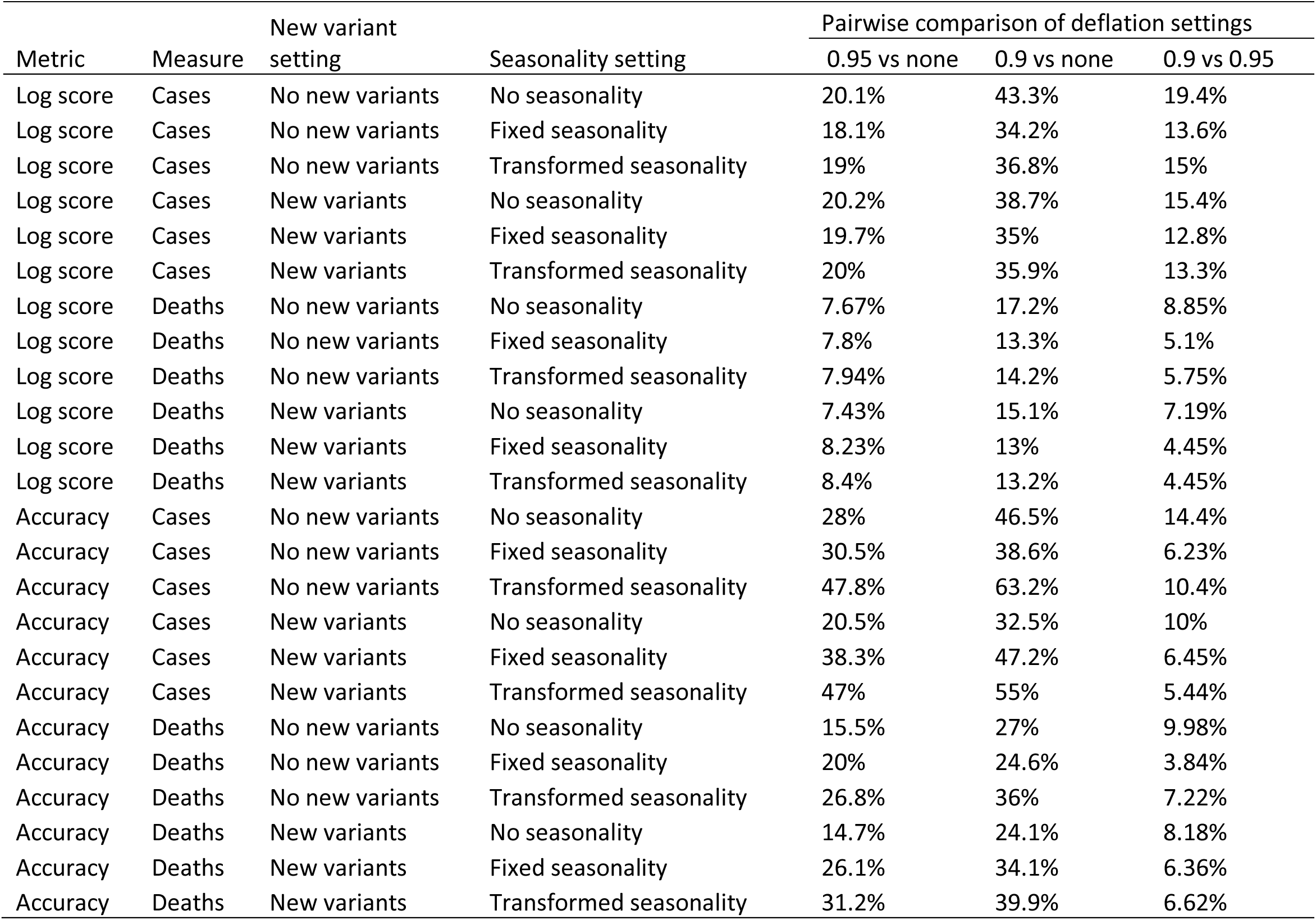
Impact of deflation. Numbers show the relative difference in mean log score computed using Eqn 6, or relative difference in mean point prediction accuracy computed using Eqn 7. For each pairwise comparison (e.g., 0.95 vs none), a positive difference indicates the former approach (e.g., 0.95) outperforms the latter (e.g., none).

| Metric | Measure | New variant setting | Seasonality setting | Pairwise comparison of deflation settings |  |  |
| --- | --- | --- | --- | --- | --- | --- |
|  |  |  |  | 0.95 vs none | 0.9 vs none | 0.9 vs 0.95 |
| Log score | Cases | No new variants | No seasonality | 20.1% | 43.3% | 19.4% |
| Log score | Cases | No new variants | Fixed seasonality | 18.1% | 34.2% | 13.6% |
| Log score | Cases | No new variants | Transformed seasonality | 19% | 36.8% | 15% |
| Log score | Cases | New variants | No seasonality | 20.2% | 38.7% | 15.4% |
| Log score | Cases | New variants | Fixed seasonality | 19.7% | 35% | 12.8% |
| Log score | Cases | New variants | Transformed seasonality | 20% | 35.9% | 13.3% |
| Log score | Deaths | No new variants | No seasonality | 7.67% | 17.2% | 8.85% |
| Log score | Deaths | No new variants | Fixed seasonality | 7.8% | 13.3% | 5.1% |
| Log score | Deaths | No new variants | Transformed seasonality | 7.94% | 14.2% | 5.75% |
| Log score | Deaths | New variants | No seasonality | 7.43% | 15.1% | 7.19% |
| Log score | Deaths | New variants | Fixed seasonality | 8.23% | 13% | 4.45% |
| Log score | Deaths | New variants | Transformed seasonality | 8.4% | 13.2% | 4.45% |
| Accuracy | Cases | No new variants | No seasonality | 28% | 46.5% | 14.4% |
| Accuracy | Cases | No new variants | Fixed seasonality | 30.5% | 38.6% | 6.23% |
| Accuracy | Cases | No new variants | Transformed seasonality | 47.8% | 63.2% | 10.4% |
| Accuracy | Cases | New variants | No seasonality | 20.5% | 32.5% | 10% |
| Accuracy | Cases | New variants | Fixed seasonality | 38.3% | 47.2% | 6.45% |
| Accuracy | Cases | New variants | Transformed seasonality | 47% | 55% | 5.44% |
| Accuracy | Deaths | No new variants | No seasonality | 15.5% | 27% | 9.98% |
| Accuracy | Deaths | No new variants | Fixed seasonality | 20% | 24.6% | 3.84% |
| Accuracy | Deaths | No new variants | Transformed seasonality | 26.8% | 36% | 7.22% |
| Accuracy | Deaths | New variants | No seasonality | 14.7% | 24.1% | 8.18% |
| Accuracy | Deaths | New variants | Fixed seasonality | 26.1% | 34.1% | 6.36% |
| Accuracy | Deaths | New variants | Transformed seasonality | 31.2% | 39.9% | 6.62% |

**Table S2.** Impact of new variants settings. Numbers show the relative difference in mean log score computed using Eqn 6, or relative difference in mean point prediction accuracy computed using Eqn 7, by variant wave. A positive number indicates superior performance of the forecast approach with anticipation of new variant emergence.

| Metric | Measure | Seasonality setting | 2nd wave | Alpha | Delta | Omicron |
| --- | --- | --- | --- | --- | --- | --- |
| Log score | Cases | No seasonality | -2.09% | 0.0271% | 66.9% | 10.5% |
| Log score | Cases | Fixed seasonality | -0.573% | -0.33% | 119% | 36.6% |
| Log score | Cases | Transformed seasonality | -1.07% | -1.03% | 93.7% | 34.1% |
| Log score | Deaths | No seasonality | -1.7% | 0.00306% | 19.1% | -5.31% |
| Log score | Deaths | Fixed seasonality | -0.387% | -0.291% | 34.1% | -2.27% |
| Log score | Deaths | Transformed seasonality | -0.918% | -0.651% | 27.8% | -2.73% |
| Accuracy | Cases | No seasonality | -1.52% | -0.11% | 89.1% | 37.1% |
| Accuracy | Cases | Fixed seasonality | 0.578% | 2.16% | 16.8% | 95.8% |
| Accuracy | Cases | Transformed seasonality | -1.33% | 1.96% | 26.1% | 77.8% |
| Accuracy | Deaths | No seasonality | -1.45% | 0.121% | 69.9% | 40.2% |
| Accuracy | Deaths | Fixed seasonality | -0.278% | 2.73% | 15.9% | 67.7% |
| Accuracy | Deaths | Transformed seasonality | -1.31% | 2.47% | 22.5% | 63.2% |

**Table S3.** Impact of seasonality, aggregated over all 10 states. Numbers show the relative difference in mean log score or point prediction accuracy, the median of pair-wise difference in log score (95% CI of the median); asterisk (*) indicates if the median is significantly >0 or <0 at the α = 0.05 level, per a Wilcoxon rank sum test. A positive difference indicates superior log score or point prediction accuracy of the first listed approach; a negative difference indicates superior log score or point prediction accuracy of the second listed approach.

| Wave | Season | Metric | Measure | Fixed vs no seasonality | Transformed vs no seasonality | Transformed vs fixed seasonality |
| --- | --- | --- | --- | --- | --- | --- |
| All | All | Log score | Cases | -4.45%, -0.0075 (-0.012, -0.0034)* | -2.8%, -0.006 (-0.01, -0.002)* | 1.73%, 0.01 (0, 0.01)* |
| All | All | Log score | Deaths | 12.7%, 0.04 (0.04, 0.04)* | 14.2%, 0.04 (0.04, 0.04)* | 1.32%, -0.0015 (-0.002, -0.00054)* |
| All | All | Accuracy | Cases | 20.4%, 10% (10%, 10%) | 26.3%, 10% (10%, 10%) | 4.92%, 0% (0%, 5%)* |
| All | All | Accuracy | Deaths | 15.8%, 10% (9.99%, 10%) | 18.2%, 10% (10%, 10%) | 2.09%, 0% (0%, 0%)* |
| All | Respiratory season | Log score | Cases | 39.8%, 0.19 (0.18, 0.2) | 42.3%, 0.2 (0.18, 0.21) | 1.8%, 0.01 (0, 0.01)* |
| All | Respiratory season | Log score | Deaths | 37.4%, 0.16 (0.15, 0.17) | 41.1%, 0.17 (0.15, 0.18) | 2.64%, 0.01 (0, 0.01)* |
| All | Respiratory season | Accuracy | Cases | 60.3%, 20% (20%, 25%) | 75.3%, 25% (25%, 30%) | 9.38%, 5.01% (5%, 5%) |
| All | Respiratory season | Accuracy | Deaths | 47.3%, 25% (20%, 25%) | 54.3%, 25% (25%, 30%) | 4.76%, 5% (0%, 5%) |
| All | Off season | Log score | Cases | -25.9%, -0.14 (-0.14, -0.13)* | -24.6%, -0.14 (-0.16, -0.13)* | 1.68%, 0 (0, 0.01)* |
| All | Off season | Log score | Deaths | -1.3%, 0.02 (0.02, 0.03)* | -0.86%, 0.02 (0.02, 0.02)* | 0.444%, -0.0031 (-0.0036, -0.0025)* |
| All | Off season | Accuracy | Cases | -5.95%, -5% (-5%, 0%)* | -6.04%, 0% (-4.99%, 0%)* | -0.0987%, 0% (0%, 0.01%)* |
| All | Off season | Accuracy | Deaths | -6.77%, -5% (-5%, -4.99%)* | -7.64%, -4.99% (-5%, -5%)* | -0.94%, 0% (0%, 0.01%)* |
| 2nd wave | Off season | Log score | Cases | 32%, 0.22 (0.2, 0.24) | 23.5%, 0.16 (0.14, 0.17) | -6.42%, -0.056 (-0.063, -0.051)* |
| 2nd wave | Off season | Log score | Deaths | 23.9%, 0.17 (0.16, 0.19) | 17.1%, 0.13 (0.12, 0.14) | -5.5%, -0.048 (-0.052, -0.043)* |
| 2nd wave | Off season | Accuracy | Cases | 37.6%, 15% (15%, 20%) | 35.6%, 20% (15%, 20%) | -1.43%, -0.01% (-5%, 0%)* |
| 2nd wave | Off season | Accuracy | Deaths | 45.6%, 25% (20%, 30%) | 28.1%, 20% (15%, 20%) | -12%, -10% (-15%, -10%)* |
| Alpha | Off season | Log score | Cases | -9.35%, 0.01 (0, 0.01)* | -24.7%, -0.052 (-0.11, 0)* | -16.9%, -0.087 (-0.11, -0.052)* |
| Alpha | Off season | Log score | Deaths | -3.33%, 0.01 (0.01, 0.01)* | -10.8%, 0.01 (0.01, 0.02)* | -7.74%, 0 (0, 0)* |
| Alpha | Off season | Accuracy | Cases | -42.4%, -30% (-35%, -25%)* | -39.8%, -30% (-35%, -25%)* | 4.52%, 5% (-0.01%, 5.01%) |
| Alpha | Off season | Accuracy | Deaths | -28.5%, -25% (-25%, -20%)* | -33.8%, -30% (-30%, -25%)* | -7.53%, -5.01% (-10%, 0%)* |
| Delta | Off season | Log score | Cases | -49.6%, -0.57 (-0.61, -0.55)* | -35.4%, -0.32 (-0.35, -0.3)* | 28.2%, 0.25 (0.23, 0.26) |
| Delta | Off season | Log score | Deaths | -12%, 0.02 (0.02, 0.02)* | -6.52%, 0.02 (0.02, 0.02)* | 6.28%, -0.0024 (-0.003, -0.0015)* |
| Delta | Off season | Accuracy | Cases | -55.8%, -25% (-25%, -20%)* | -50%, -20% (-25%, -20%)* | 13.1%, 4.99% (0%, 5%)* |
| Delta | Off season | Accuracy | Deaths | -50.3%, -25% (-25%, -20%)* | -43.2%, -20% (-20%, -15%)* | 14.5%, 5% (4.99%, 10%) |
| Omicron | Off season | Log score | Cases | -19.5%, -0.15 (-0.16, -0.14)* | -24%, -0.16 (-0.17, -0.15)* | -5.68%, -0.013 (-0.017, -0.0085)* |
| Omicron | Off season | Log score | Deaths | 4.77%, 0.03 (0.03, 0.03)* | 3.84%, 0.02 (0.02, 0.03)* | -0.887%, -0.0035 (-0.0041, -0.003)* |
| Omicron | Off season | Accuracy | Cases | 4.46%, 0.01% (0%, 5%)* | 2.52%, 0% (0%, 5%)* | -1.86%, -0.01% (0%, 0%)* |
| Omicron | Off season | Accuracy | Deaths | -1.59%, 0% (0%, 0%)* | -1.78%, 0% (0%, 0%)* | -0.191%, -0.01% (0%, 0%)* |

**Table S4.** Impact of seasonality, by state. Numbers show the relative difference in mean log score or point prediction accuracy, the median of pair-wise difference in log score (95% CI of the median); asterisk (*) indicates if the median is significantly >0 or <0 at the α = 0.05 level, per a Wilcoxon rank sum test. A positive difference indicates superior log score or point prediction accuracy of the first listed approach; a negative difference indicates superior log score or point prediction accuracy of the second listed approach.

| State | Season | Metric | Measure | Fixed vs no seasonality | Transformed vs no seasonality | Transformed vs fixed seasonality |
| --- | --- | --- | --- | --- | --- | --- |
| California | All | Log score | Cases | -7.14%, -0.042 (-0.062, -0.024)* | -1.53%, -0.028 (-0.045, -0.013)* | 6.04%, 0.02 (0.01, 0.02)* |
| California | All | Log score | Deaths | 16.7%, 0.04 (0.04, 0.05)* | 21.4%, 0.04 (0.03, 0.04)* | 3.96%, 0 (0, 0.01)* |
| California | All | Accuracy | Cases | 13.1%, 5% (0%, 9.99%) | 11.3%, 5% (0%, 10%) | -1.6%, 0% (0%, 0%)* |
| California | All | Accuracy | Deaths | 4.74%, 0% (0.01%, 5%)* | 1.86%, 0% (-0.01%, 5%)* | -2.75%, 0% (-4.99%, 0%)* |
| California | Respiratory season | Log score | Cases | 42.5%, 0.17 (0.13, 0.22) | 56.7%, 0.24 (0.19, 0.3) | 9.96%, 0.05 (0.04, 0.06)* |
| California | Respiratory season | Log score | Deaths | 48.4%, 0.21 (0.15, 0.3) | 60%, 0.27 (0.19, 0.37) | 7.8%, 0.03 (0.02, 0.04)* |
| California | Respiratory season | Accuracy | Cases | 51.9%, 20% (15%, 20%) | 58.9%, 20% (15%, 25%) | 4.61%, 4.99% (-0.01%, 5%)* |
| California | Respiratory season | Accuracy | Deaths | 27.9%, 15% (10%, 20%) | 22.4%, 10% (5%, 15%) | -4.33%, -5% (-10%, 0%)* |
| California | Off season | Log score | Cases | -30.3%, -0.17 (-0.2, -0.14)* | -27.9%, -0.19 (-0.23, -0.16)* | 3.49%, -0.0024 (-0.013, 0)* |
| California | Off season | Log score | Deaths | -0.598%, 0.03 (0.02, 0.03)* | 0.856%, 0.02 (0.02, 0.03)* | 1.46%, -1e-05 (-0.0014, 0.00098)* |
| California | Off season | Accuracy | Cases | -11%, -5% (-10%, 0%)* | -18.4%, -9.99% (-15%, -5.01%)* | -8.21%, -5% (-10%, 0%)* |
| California | Off season | Accuracy | Deaths | -8.26%, -5% (-10%, 0%)* | -9.65%, -5% (-10%, -5%)* | -1.52%, 0% (-4.99%, 5%)* |
| Florida | All | Log score | Cases | -12.6%, -0.02 (-0.032, -0.01)* | -6.11%, 0.001 (-0.0085, 0.01)* | 7.47%, 0.03 (0.02, 0.04)* |
| Florida | All | Log score | Deaths | 0.697%, 0.02 (0.01, 0.02)* | 4.42%, 0.02 (0.02, 0.03)* | 3.7%, 0.01 (0.01, 0.01)* |
| Florida | All | Accuracy | Cases | 8.52%, 5% (5%, 10%) | 11.5%, 5% (5%, 10%) | 2.74%, 0% (0%, 5%)* |
| Florida | All | Accuracy | Deaths | 3.73%, 5% (0%, 5.01%) | 4.9%, 5% (0%, 5.01%) | 1.14%, 0% (0%, 0%)* |
| Florida | Respiratory season | Log score | Cases | -2.01%, 0.03 (0.02, 0.05)* | 1.72%, 0.04 (0.02, 0.05)* | 3.81%, 0.02 (0.01, 0.04)* |
| Florida | Respiratory season | Log score | Deaths | 2.77%, 0.05 (0.04, 0.06) | 8.52%, 0.06 (0.04, 0.07) | 5.59%, 0.03 (0.02, 0.04)* |
| Florida | Respiratory season | Accuracy | Cases | 19%, 10% (9.99%, 15%) | 20.7%, 15% (10%, 20%) | 1.44%, 0% (-5%, 5%)* |
| Florida | Respiratory season | Accuracy | Deaths | 11.8%, 10% (9.99%, 15%) | 12.9%, 10% (5%, 15%) | 0.958%, 0% (-5%, 4.99%)* |
| Florida | Off season | Log score | Cases | -19.1%, -0.066 (-0.088, -0.046)* | -11%, -0.02 (-0.037, -0.0059)* | 10%, 0.03 (0.02, 0.05)* |
| Florida | Off season | Log score | Deaths | -0.675%, 0.01 (0.01, 0.01)* | 1.75%, 0.01 (0.01, 0.02)* | 2.44%, 0 (0.00049, 0)* |
| Florida | Off season | Accuracy | Cases | 2.1%, 0% (0%, 5%)* | 5.84%, 5% (0%, 5%) | 3.66%, 0% (0.01%, 5%)* |
| Florida | Off season | Accuracy | Deaths | -1.82%, 0% (-5%, 0%)* | -0.565%, 0% (0%, 4.99%)* | 1.28%, 0.01% (0%, 4.99%)* |
| Iowa | All | Log score | Cases | 1.02%, 0.02 (0.01, 0.03)* | 6.96%, 0.03 (0.02, 0.04)* | 5.89%, 0.02 (0.01, 0.03)* |
| Iowa | All | Log score | Deaths | 23.6%, 0.07 (0.06, 0.08) | 25.7%, 0.06 (0.05, 0.07) | 1.64%, -0.001 (-0.0031, 5e-04)* |
| Iowa | All | Accuracy | Cases | 12.9%, 5% (0%, 10%) | 28.3%, 10% (10%, 15%) | 13.7%, 10% (5%, 10%) |
| Iowa | All | Accuracy | Deaths | 17.8%, 10% (5%, 10%) | 22.7%, 10% (10%, 15%) | 4.19%, 5% (0.01%, 5.01%) |
| Iowa | Respiratory season | Log score | Cases | 77.8%, 0.47 (0.4, 0.53) | 77.4%, 0.48 (0.42, 0.54) | -0.246%, 0.01 (-0.003, 0.02)* |
| Iowa | Respiratory season | Log score | Deaths | 72.9%, 0.38 (0.31, 0.46) | 76.2%, 0.44 (0.36, 0.5) | 1.95%, 0.01 (-0.001, 0.02)* |
| Iowa | Respiratory season | Accuracy | Cases | 71.5%, 25% (20%, 25%) | 112%, 35% (30%, 40%) | 23.4%, 15% (9.99%, 15%) |
| Iowa | Respiratory season | Accuracy | Deaths | 81%, 35% (30%, 35%) | 88.7%, 35% (30%, 40%) | 4.26%, 5% (0.01%, 10%) |
| Iowa | Off season | Log score | Cases | -30.2%, -0.11 (-0.14, -0.076)* | -23.2%, -0.076 (-0.1, -0.05)* | 10.1%, 0.02 (0.01, 0.03)* |
| Iowa | Off season | Log score | Deaths | -0.723%, 0.03 (0.03, 0.04)* | 0.71%, 0.03 (0.03, 0.03)* | 1.44%, -0.0035 (-0.0055, -0.0025)* |
| Iowa | Off season | Accuracy | Cases | -17.6%, -10% (-15%, -5%)* | -14.9%, -10% (-10%, -4.99%)* | 3.29%, 0% (0%, 5%)* |
| Iowa | Off season | Accuracy | Deaths | -18.8%, -10% (-15%, -10%)* | -15.5%, -10% (-15%, -5.01%)* | 4.1%, 0% (0%, 5%)* |
| Massachusetts | All | Log score | Cases | 8.31%, 0.05 (0.03, 0.07) | 4.64%, 0.02 (0, 0.03)* | -3.39%, -0.03 (-0.038, -0.023)* |
| Massachusetts | All | Log score | Deaths | 30%, 0.11 (0.1, 0.12) | 28.8%, 0.08 (0.07, 0.09) | -0.937%, -0.015 (-0.018, -0.012)* |
| Massachusetts | All | Accuracy | Cases | 49.1%, 20% (15%, 25%) | 33.5%, 15% (10%, 15%) | -10.4%, -10% (-10%, -5%)* |
| Massachusetts | All | Accuracy | Deaths | 34.7%, 20% (15%, 20%) | 28.1%, 15% (10%, 20%) | -4.87%, -5% (-5%, 0%)* |
| Massachusetts | Respiratory season | Log score | Cases | 83.6%, 0.5 (0.44, 0.57) | 78%, 0.45 (0.39, 0.52) | -3.05%, -0.034 (-0.044, -0.023)* |
| Massachusetts | Respiratory season | Log score | Deaths | 72.7%, 0.42 (0.34, 0.49) | 71%, 0.41 (0.35, 0.48) | -0.976%, -0.021 (-0.028, -0.015)* |
| Massachusetts | Respiratory season | Accuracy | Cases | 121%, 35% (35%, 40%) | 99.7%, 35% (30%, 40%) | -9.73%, -10% (-10%, -5%)* |
| Massachusetts | Respiratory season | Accuracy | Deaths | 68.3%, 30% (25%, 35%) | 56.7%, 25% (25%, 30%) | -6.9%, -5% (-10%, -5%)* |
| Massachusetts | Off season | Log score | Cases | -24%, -0.074 (-0.11, -0.046)* | -26.8%, -0.18 (-0.24, -0.14)* | -3.62%, -0.026 (-0.036, -0.017)* |
| Massachusetts | Off season | Log score | Deaths | 7.39%, 0.05 (0.04, 0.06)* | 6.42%, 0.03 (0.03, 0.04)* | -0.91%, -0.012 (-0.014, -0.0096)* |
| Massachusetts | Off season | Accuracy | Cases | 3.66%, 0% (0%, 5%)* | -8.13%, -4.99% (-10%, 0%)* | -11.4%, -5% (-10%, -5%)* |
| Massachusetts | Off season | Accuracy | Deaths | 7.54%, 5% (0.01%, 9.99%) | 5.05%, 5% (0%, 5%) | -2.31%, 0% (-5%, 0%)* |
| Michigan | All | Log score | Cases | 0.571%, -0.0055 (-0.024, 0.01)* | -9.27%, -0.034 (-0.054, -0.015)* | -9.79%, -0.03 (-0.038, -0.023)* |
| Michigan | All | Log score | Deaths | 16.8%, 0.06 (0.05, 0.08) | 7.52%, 0.04 (0.03, 0.05)* | -7.95%, -0.032 (-0.036, -0.028)* |
| Michigan | All | Accuracy | Cases | 26.8%, 10% (9.99%, 15%) | 20.5%, 10% (5%, 10%) | -5.01%, -5% (-5%, 0%)* |
| Michigan | All | Accuracy | Deaths | 21.6%, 10% (9.99%, 15%) | 10.2%, 5% (5%, 9.99%) | -9.42%, -5% (-9.99%, -5%)* |
| Michigan | Respiratory season | Log score | Cases | 38.8%, 0.24 (0.19, 0.29) | 29.1%, 0.19 (0.15, 0.24) | -7.03%, -0.036 (-0.044, -0.027)* |
| Michigan | Respiratory season | Log score | Deaths | 38.7%, 0.19 (0.15, 0.24) | 33%, 0.16 (0.12, 0.2) | -4.09%, -0.036 (-0.043, -0.028)* |
| Michigan | Respiratory season | Accuracy | Cases | 116%, 35% (30%, 40%) | 84.3%, 25% (25%, 30%) | -14.6%, -10% (-15%, -5%)* |
| Michigan | Respiratory season | Accuracy | Deaths | 83.9%, 40% (35%, 40%) | 44.1%, 25% (20%, 25%) | -21.6%, -20% (-20%, -15%)* |
| Michigan | Off season | Log score | Cases | -18.6%, -0.14 (-0.18, -0.11)* | -28%, -0.21 (-0.24, -0.16)* | -11.6%, -0.029 (-0.042, -0.017)* |
| Michigan | Off season | Log score | Deaths | 4.38%, 0.04 (0.03, 0.05)* | -6.47%, 0.02 (0.01, 0.03)* | -10.4%, -0.028 (-0.032, -0.024)* |
| Michigan | Off season | Accuracy | Cases | -16.4%, -10% (-10%, -5%)* | -10.6%, -5.01% (-9.99%, 0%)* | 7.01%, 5% (0%, 10%) |
| Michigan | Off season | Accuracy | Deaths | -18%, -10% (-15%, -10%)* | -11.4%, -5% (-10%, 0%)* | 8.01%, 5.01% (0%, 10%) |
| New York | All | Log score | Cases | -3.26%, 0.01 (5.9e-06, 0.02)* | -4.56%, -0.002 (-0.02, 0.01)* | -1.34%, -0.0065 (-0.014, 6.4e-05)* |
| New York | All | Log score | Deaths | 19%, 0.08 (0.07, 0.09) | 17.9%, 0.06 (0.05, 0.07) | -0.852%, -0.011 (-0.014, -0.0086)* |
| New York | All | Accuracy | Cases | 30.7%, 15% (10%, 15%) | 51%, 20% (20%, 25%) | 15.6%, 10% (5%, 15%) |
| New York | All | Accuracy | Deaths | 21.7%, 10% (10%, 15%) | 32.1%, 20% (15%, 20%) | 8.58%, 10% (5%, 10%) |
| New York | Respiratory season | Log score | Cases | 44.9%, 0.24 (0.21, 0.28) | 44.3%, 0.24 (0.2, 0.28) | -0.435%, -0.007 (-0.019, 0)* |
| New York | Respiratory season | Log score | Deaths | 37.8%, 0.21 (0.18, 0.25) | 37.8%, 0.21 (0.17, 0.24) | -0.00206%, -0.0056 (-0.014, 0)* |
| New York | Respiratory season | Accuracy | Cases | 62.7%, 25% (20%, 30%) | 100%, 35% (30%, 40%) | 22.9%, 15% (10%, 20%) |
| New York | Respiratory season | Accuracy | Deaths | 51.3%, 25% (20%, 30%) | 80.5%, 40% (35%, 40%) | 19.3%, 15% (15%, 20%) |
| New York | Off season | Log score | Cases | -26.3%, -0.11 (-0.14, -0.085)* | -27.7%, -0.2 (-0.25, -0.15)* | -1.94%, -0.0074 (-0.017, 2.2e-05)* |
| New York | Off season | Log score | Deaths | 7.78%, 0.05 (0.05, 0.06) | 6.25%, 0.04 (0.03, 0.04)* | -1.42%, -0.012 (-0.014, -0.01)* |
| New York | Off season | Accuracy | Cases | 6.51%, 5% (-0.01%, 10%) | 14%, 5% (0%, 10%) | 7.02%, 5% (0%, 10%) |
| New York | Off season | Accuracy | Deaths | -0.706%, -0.01% (-4.99%, 5%)* | -4.44%, -5% (-10%, 0%)* | -3.76%, -5% (-5%, 0%)* |
| Pennsylvania | All | Log score | Cases | -5.3%, -0.008 (-0.026, 0.01)* | -5.72%, 0.01 (-0.0085, 0.02)* | -0.438%, -0.0039 (-0.011, 0)* |
| Pennsylvania | All | Log score | Deaths | 13.8%, 0.07 (0.06, 0.08) | 12.9%, 0.06 (0.05, 0.07) | -0.791%, -0.012 (-0.016, -0.009)* |
| Pennsylvania | All | Accuracy | Cases | 29.7%, 10% (10%, 15%) | 47.5%, 20% (15%, 20%) | 13.7%, 10% (5%, 10%) |
| Pennsylvania | All | Accuracy | Deaths | 29.4%, 15% (10%, 20%) | 36.4%, 20% (15%, 20%) | 5.39%, 5% (0%, 9.99%) |
| Pennsylvania | Respiratory season | Log score | Cases | 38.5%, 0.18 (0.14, 0.22) | 38.6%, 0.2 (0.17, 0.23) | 0.0339%, -0.003 (-0.016, 0.01)* |
| Pennsylvania | Respiratory season | Log score | Deaths | 36.2%, 0.15 (0.12, 0.18) | 35.4%, 0.14 (0.11, 0.16) | -0.636%, -0.017 (-0.028, -0.006)* |
| Pennsylvania | Respiratory season | Accuracy | Cases | 48.9%, 20% (15%, 25%) | 99.6%, 35% (30%, 40%) | 34.1%, 20% (15%, 25%) |
| Pennsylvania | Respiratory season | Accuracy | Deaths | 50.6%, 25% (20%, 30%) | 76.7%, 40% (35%, 40%) | 17.4%, 15% (10%, 20%) |
| Pennsylvania | Off season | Log score | Cases | -26.7%, -0.18 (-0.23, -0.14)* | -27.2%, -0.19 (-0.24, -0.14)* | -0.754%, -0.0046 (-0.012, 0)* |
| Pennsylvania | Off season | Log score | Deaths | 0.845%, 0.05 (0.04, 0.06)* | -0.0572%, 0.04 (0.04, 0.05)* | -0.895%, -0.011 (-0.014, -0.0085)* |
| Pennsylvania | Off season | Accuracy | Cases | 13%, 5% (0.01%, 9.99%) | 2.05%, 0% (-0.01%, 4.99%)* | -9.71%, -5.01% (-10%, -4.99%)* |
| Pennsylvania | Off season | Accuracy | Deaths | 11.1%, 5% (0%, 10%) | 1.46%, 0% (-4.99%, 5%)* | -8.66%, -5% (-10%, -5%)* |
| Texas | All | Log score | Cases | -9.81%, -0.02 (-0.043, -0.00054)* | 1.82%, 0.01 (-0.0066, 0.02)* | 12.9%, 0.06 (0.05, 0.07) |
| Texas | All | Log score | Deaths | 6.34%, 0.03 (0.02, 0.03)* | 15.6%, 0.03 (0.02, 0.03)* | 8.68%, 0.01 (0.01, 0.01)* |
| Texas | All | Accuracy | Cases | 11.1%, 5% (0.01%, 5%) | 33.5%, 15% (10%, 15%) | 20.2%, 10% (10%, 15%) |
| Texas | All | Accuracy | Deaths | 9.13%, 5% (0%, 5%) | 27.6%, 15% (10%, 15%) | 16.9%, 10% (9.99%, 15%) |
| Texas | Respiratory season | Log score | Cases | 59.4%, 0.34 (0.26, 0.42) | 77.3%, 0.43 (0.36, 0.51) | 11.3%, 0.05 (0.04, 0.07) |
| Texas | Respiratory season | Log score | Deaths | 51.9%, 0.3 (0.23, 0.37) | 68%, 0.42 (0.35, 0.49) | 10.6%, 0.07 (0.05, 0.08) |
| Texas | Respiratory season | Accuracy | Cases | 50.1%, 20% (15%, 25%) | 96.9%, 30% (25%, 35%) | 31.2%, 20% (15%, 25%) |
| Texas | Respiratory season | Accuracy | Deaths | 47%, 25% (20%, 25%) | 91.5%, 40% (35%, 40%) | 30.2%, 25% (20%, 25%) |
| Texas | Off season | Log score | Cases | -38.5%, -0.22 (-0.27, -0.18)* | -29.9%, -0.13 (-0.17, -0.096)* | 14%, 0.06 (0.05, 0.08) |
| Texas | Off season | Log score | Deaths | -16.3%, 0.02 (0.02, 0.02)* | -10.1%, 0.01 (0.01, 0.02)* | 7.43%, -1.8e-06 (-0.00099, 0.00049)* |
| Texas | Off season | Accuracy | Cases | -16%, -10% (-15%, -5.01%)* | -10.5%, -5% (-10%, -5%)* | 6.57%, 5% (0.01%, 5%) |
| Texas | Off season | Accuracy | Deaths | -18.8%, -15% (-15%, -10%)* | -19.6%, -15% (-15%, -10%)* | -0.909%, 0% (-0.01%, 0%)* |
| Washington | All | Log score | Cases | -14%, -0.092 (-0.11, -0.078)* | -13.5%, -0.098 (-0.11, -0.084)* | 0.577%, 0.01 (0, 0.01)* |
| Washington | All | Log score | Deaths | -4.44%, -0.002 (-0.007, 0.00092)* | -1.65%, 0 (-0.0015, 0)* | 2.92%, 0.01 (0.01, 0.01)* |
| Washington | All | Accuracy | Cases | 16.5%, 5% (4.99%, 10%) | 13.5%, 5% (0.01%, 10%) | -2.54%, 0% (0%, 0.01%)* |
| Washington | All | Accuracy | Deaths | 11%, 5.01% (0%, 10%) | 11.3%, 5.01% (5%, 9.99%) | 0.291%, 0% (-0.01%, 0%)* |
| Washington | Respiratory season | Log score | Cases | 0.837%, -0.026 (-0.046, -0.0079)* | 3.75%, -0.0025 (-0.018, 0.01)* | 2.89%, 0.03 (0.02, 0.03)* |
| Washington | Respiratory season | Log score | Deaths | 0.291%, -0.05 (-0.068, -0.032)* | 2.95%, -0.032 (-0.046, -0.017)* | 2.65%, 0.02 (0.01, 0.02)* |
| Washington | Respiratory season | Accuracy | Cases | 27%, 10% (5%, 15%) | 25.6%, 10% (5.01%, 15%) | -1.03%, 0% (-0.01%, 0%)* |
| Washington | Respiratory season | Accuracy | Deaths | 23.9%, 10% (10%, 15%) | 29.2%, 15% (10%, 20%) | 4.31%, 4.99% (0%, 5%)* |
| Washington | Off season | Log score | Cases | -22.7%, -0.15 (-0.17, -0.13)* | -23.4%, -0.19 (-0.22, -0.17)* | -0.949%, -0.01 (-0.018, -0.0035)* |
| Washington | Off season | Log score | Deaths | -7.49%, 0.01 (0.01, 0.01)* | -4.62%, 0.01 (0.01, 0.01)* | 3.1%, 0 (0, 0)* |
| Washington | Off season | Accuracy | Cases | 5.51%, 0.01% (0%, 5.01%)* | 0.819%, 0.01% (0%, 5%)* | -4.45%, 0% (-5%, 0%)* |
| Washington | Off season | Accuracy | Deaths | -1.29%, -0.01% (-5%, 0%)* | -5.72%, -5% (-5%, 0%)* | -4.5%, -5% (-5%, 0%)* |
| Wyoming | All | Log score | Cases | 0.44%, -0.00092 (-0.007, 0)* | 1.31%, 0 (-0.0045, 0.01)* | 0.869%, 0.01 (0.01, 0.01)* |
| Wyoming | All | Log score | Deaths | 8.84%, 0.03 (0.02, 0.03)* | 12.6%, 0.03 (0.02, 0.03)* | 3.47%, 0 (0, 0.01)* |
| Wyoming | All | Accuracy | Cases | 6.6%, 4.99% (0%, 5%)* | 14.4%, 5% (5%, 10%) | 7.28%, 5% (0.01%, 5.01%) |
| Wyoming | All | Accuracy | Deaths | 3.37%, 0.01% (0%, 5%)* | 7.98%, 5% (0%, 5%) | 4.46%, 0% (-0.01%, 5%)* |
| Wyoming | Respiratory season | Log score | Cases | 40.7%, 0.19 (0.16, 0.22) | 42.9%, 0.17 (0.15, 0.2) | 1.51%, -0.0036 (-0.013, 0)* |
| Wyoming | Respiratory season | Log score | Deaths | 34.1%, 0.18 (0.14, 0.21) | 39.6%, 0.16 (0.13, 0.19) | 4.1%, 8.3e-05 (-0.0084, 0.01)* |
| Wyoming | Respiratory season | Accuracy | Cases | 57.9%, 20% (15%, 25%) | 70.8%, 25% (20%, 30%) | 8.14%, 5% (0.01%, 10%) |
| Wyoming | Respiratory season | Accuracy | Deaths | 29.6%, 15% (10%, 20%) | 41.1%, 20% (15%, 25%) | 8.87%, 5% (0%, 10%) |
| Wyoming | Off season | Log score | Cases | -19.5%, -0.15 (-0.18, -0.12)* | -19.1%, -0.13 (-0.16, -0.093)* | 0.454%, 0.02 (0.01, 0.02)* |
| Wyoming | Off season | Log score | Deaths | -5.06%, 0.01 (0.01, 0.01)* | -2.16%, 0.01 (0.01, 0.02)* | 3.05%, 0.01 (0, 0.01)* |
| Wyoming | Off season | Accuracy | Cases | -20%, -10% (-15%, -5%)* | -14.9%, -5% (-10%, 0%)* | 6.39%, 5% (0%, 10%) |
| Wyoming | Off season | Accuracy | Deaths | -14.2%, -9.99% (-10%, -5.01%)* | -14.2%, -9.99% (-10%, -5%)* | 0%, 0% (-0.01%, 5%)* |

**Table S5.** Comparison of forecast performance of the ARIMAX models. Only four models (see the top row for model names) are shown here because the fifth model (ARIMAX.FULL with vaccination included) was only able to generate forecasts for less than half of the study weeks; see details on the models in the main text. Numbers show the mean log score or point prediction accuracy of forecasts (specified in the “metric” column), aggregated across the entire study period and all locations for all forecast targets combined or individual forecast targets (specified in the “target” column). Bolded fonts indicate best performance (highest log score or accuracy).

| target | metric | measure | Models |  |  |  |
| --- | --- | --- | --- | --- | --- | --- |
|  |  |  | ARIMA | ARIMAX.MOB | ARIMAX.SN | ARIMAX.MS |
| all | Log score | Cases | -2.64 | **-2.53** | -2.75 | -3.03 |
| all | Log score | Deaths | -1.77 | -1.73 | **-1.64** | -1.71 |
| all | Accuracy | Cases | 13% | 15% | **18%** | 15% |
| all | Accuracy | Deaths | 14% | 17% | **21%** | 17% |
| 1-8wk ahead | Log score | Cases | -2.08 | **-2** | -2.04 | -2.08 |
| 1-8wk ahead | Log score | Deaths | -1.34 | -1.36 | **-1.27** | -1.34 |
| 1-8wk ahead | Accuracy | Cases | 22% | 25% | **26%** | 24% |
| 1-8wk ahead | Accuracy | Deaths | 22% | 25% | **28%** | 23% |
| 9-16wk ahead | Log score | Cases | -2.86 | **-2.62** | -2.8 | -3.14 |
| 9-16wk ahead | Log score | Deaths | -1.89 | -1.8 | **-1.64** | -1.68 |
| 9-16wk ahead | Accuracy | Cases | 8% | 11% | **14%** | 11% |
| 9-16wk ahead | Accuracy | Deaths | 8% | 12% | **18%** | 12% |
| 17-26wk ahead | Log score | Cases | -2.89 | **-2.77** | -3.26 | -3.77 |
| 17-26wk ahead | Log score | Deaths | -1.82 | -1.75 | **-1.72** | -1.76 |
| 17-26wk ahead | Accuracy | Cases | 8% | 10% | **12%** | 10% |
| 17-26wk ahead | Accuracy | Deaths | 10% | 12% | **16%** | 12% |
| peak intensity | Log score | Cases | **-3.87** | -4.45 | -4.39 | -4.57 |
| peak intensity | Log score | Deaths | **-3.14** | -3.38 | -3.18 | -3.5 |
| peak intensity | Accuracy | Cases | 14% | 15% | **19%** | 18% |
| peak intensity | Accuracy | Deaths | 15% | 18% | **23%** | 21% |
| peak week | Log score | Cases | **-2.63** | -2.66 | -3.01 | -3.21 |
| peak week | Log score | Deaths | -2.54 | -2.53 | **-2.36** | -2.45 |
| peak week | Accuracy | Cases | 11% | 12% | **20%** | 21% |
| peak week | Accuracy | Deaths | 11% | 13% | **23%** | 23% |
| total | Log score | Cases | -2.37 | **-2.36** | -2.54 | -2.5 |
| total | Log score | Deaths | **-2.22** | -2.27 | -2.29 | -2.56 |
| total | Accuracy | Cases | 10% | 12% | **13%** | **13%** |
| total | Accuracy | Deaths | 12% | 13% | 17% | **19%** |

**Table S6.** Comparison of forecast performance of the approaches developed in this study with the best-performing ARIMAX model. Numbers show the mean log score or point prediction accuracy of forecasts (specified in the “metric” column), aggregated across the entire study period and all locations for all forecast targets combined or individual forecast targets (specified in the “target” column). Bolded fonts indicate best performance (highest log score or accuracy).

| target | measure | Log score |  |  | Accuracy |  |  |
| --- | --- | --- | --- | --- | --- | --- | --- |
|  |  | ARIMAX.SN | Baseline | Best-performing | ARIMAX.SN | Baseline | Best-performing |
| all | Cases | -2.75 | -1.95 | **-1.46** | 18% | 11% | **26%** |
| all | Deaths | -1.64 | -0.97 | **-0.65** | 21% | 17% | **31%** |
| 1-8wk ahead | Cases | -2.04 | -1.08 | **-0.91** | 26% | 26% | **38%** |
| 1-8wk ahead | Deaths | -1.27 | -0.42 | **-0.35** | 28% | 39% | **48%** |
| 9-16wk ahead | Cases | -2.8 | -1.86 | **-1.49** | 14% | 4% | **20%** |
| 9-16wk ahead | Deaths | -1.64 | -0.83 | **-0.64** | 18% | 7% | **25%** |
| 17-26wk ahead | Cases | -3.26 | -2.8 | **-1.87** | 12% | 1% | **16%** |
| 17-26wk ahead | Deaths | -1.72 | -1.43 | **-0.8** | **16%** | 1% | **16%** |
| peak intensity | Cases | -4.39 | -2.43 | **-2.01** | 19% | 20% | **40%** |
| peak intensity | Deaths | -3.18 | -1.69 | **-1.36** | 23% | 30% | **51%** |
| peak week | Cases | -3.01 | -3.51 | **-2.73** | 20% | 24% | **42%** |
| peak week | Deaths | -2.36 | -2.61 | **-1.53** | 23% | 35% | **56%** |
| total | Cases | -2.54 | -1.05 | **-0.75** | 13% | 7% | **33%** |
| total | Deaths | -2.29 | -0.99 | **-0.67** | 17% | 9% | **36%** |

**Table S7.** Preliminary assessment of the real-time forecasts initiated the week of October 2, 2022 for October 2022 – March 2023. The log score and accuracy were computed using reported case and mortality data downloaded on March 31, 2023 (see further details in the main text). As shown in Fig 8, COVID-19 mortality data in some states (e.g., Wyoming) were highly irregular during the forecast period, likely an artifact of reporting. Due to these potential data inaccuracies, the mortality-related log score and point prediction accuracy for these states are likely lower than the true values (to be obtained once more complete mortality data are available).

| State | target | Log score |  | Accuracy |  |
| --- | --- | --- | --- | --- | --- |
|  |  | Cases | Deaths | Cases | Deaths |
| All | all | -0.45 | -0.2 | 56% | 23% |
| All | 1-8wk ahead | -0.31 | -0.06 | 56% | 22% |
| All | 9-16wk ahead | -0.71 | -0.17 | 46% | 20% |
| All | 17-26wk ahead | -0.25 | -0.12 | 64% | 28% |
| All | peak intensity | -0.84 | -0.32 | 48% | 6% |
| All | peak week | -1.21 | -2.3 | 52% | 41% |
| All | total | -0.21 | -0.13 | 78% | 7% |
| California | all | -0.52 | -0.1 | 60% | 52% |
| California | 1-8wk ahead | -0.31 | -0.04 | 55% | 32% |
| California | 9-16wk ahead | -0.99 | -0.11 | 41% | 74% |
| California | 17-26wk ahead | -0.24 | -0.06 | 73% | 43% |
| California | peak intensity | -1.39 | -0.1 | 90% | 60% |
| California | peak week | -0.72 | -0.86 | 100% | 100% |
| California | total | -0.24 | -0.1 | 60% | 60% |
| Florida | all | -0.38 | -0.19 | 28% | 8% |
| Florida | 1-8wk ahead | -0.06 | -0.02 | 5% | 0% |
| Florida | 9-16wk ahead | -0.66 | -0.2 | 9% | 14% |
| Florida | 17-26wk ahead | -0.31 | -0.16 | 56% | 13% |
| Florida | peak intensity | -1.2 | -0.54 | 0% | 0% |
| Florida | peak week | -0.74 | -1.6 | 90% | 0% |
| Florida | total | -0.16 | -0.05 | 50% | 0% |
| Iowa | all | -0.37 | -0.12 | 53% | 29% |
| Iowa | 1-8wk ahead | -0.2 | -0.05 | 86% | 12% |
| Iowa | 9-16wk ahead | -0.59 | -0.09 | 42% | 24% |
| Iowa | 17-26wk ahead | -0.2 | -0.07 | 29% | 49% |
| Iowa | peak intensity | -0.71 | -0.11 | 80% | 0% |
| Iowa | peak week | -1.37 | -1.38 | 60% | 70% |
| Iowa | total | -0.26 | -0.05 | 70% | 0% |
| Massachusetts | all | -0.64 | -0.31 | 61% | 9% |
| Massachusetts | 1-8wk ahead | -0.7 | -0.06 | 41% | 18% |
| Massachusetts | 9-16wk ahead | -0.86 | -0.46 | 72% | 6% |
| Massachusetts | 17-26wk ahead | -0.36 | -0.2 | 78% | 0% |
| Massachusetts | peak intensity | -1.33 | -0.61 | 0% | 0% |
| Massachusetts | peak week | -0.95 | -1.62 | 0% | 70% |
| Massachusetts | total | -0.26 | -0.7 | 80% | 0% |
| Michigan | all | -0.32 | -0.32 | 51% | 17% |
| Michigan | 1-8wk ahead | -0.3 | -0.2 | 45% | 25% |
| Michigan | 9-16wk ahead | -0.46 | -0.21 | 64% | 12% |
| Michigan | 17-26wk ahead | -0.11 | -0.22 | 41% | 19% |
| Michigan | peak intensity | -0.35 | -0.12 | 100% | 0% |
| Michigan | peak week | -1.81 | -3.59 | 0% | 0% |
| Michigan | total | -0.06 | -0.05 | 90% | 0% |
| New York | all | -0.76 | -0.14 | 66% | 17% |
| New York | 1-8wk ahead | -0.68 | -0.07 | 52% | 14% |
| New York | 9-16wk ahead | -1.27 | -0.05 | 48% | 0% |
| New York | 17-26wk ahead | -0.34 | -0.05 | 98% | 37% |
| New York | peak intensity | -0.97 | -0.11 | 30% | 0% |
| New York | peak week | -1.76 | -2.31 | 10% | 0% |
| New York | total | -0.24 | -0.06 | 90% | 0% |
| Pennsylvania | all | -0.49 | -0.12 | 59% | 37% |
| Pennsylvania | 1-8wk ahead | -0.33 | -0.05 | 81% | 25% |
| Pennsylvania | 9-16wk ahead | -0.75 | -0.1 | 34% | 30% |
| Pennsylvania | 17-26wk ahead | -0.34 | -0.08 | 62% | 53% |
| Pennsylvania | peak intensity | -0.73 | -0.16 | 0% | 0% |
| Pennsylvania | peak week | -1.09 | -1.33 | 100% | 90% |
| Pennsylvania | total | -0.21 | -0.08 | 80% | 0% |
| Texas | all | -0.37 | -0.08 | 64% | 10% |
| Texas | 1-8wk ahead | -0.22 | -0.03 | 78% | 4% |
| Texas | 9-16wk ahead | -0.67 | -0.05 | 60% | 19% |
| Texas | 17-26wk ahead | -0.18 | -0.04 | 50% | 3% |
| Texas | peak intensity | -0.75 | -0.09 | 70% | 0% |
| Texas | peak week | -0.83 | -1.22 | 100% | 70% |
| Texas | total | -0.26 | -0.05 | 80% | 0% |
| Washington | all | -0.26 | -0.3 | 78% | 31% |
| Washington | 1-8wk ahead | -0.21 | -0.04 | 75% | 65% |
| Washington | 9-16wk ahead | -0.35 | -0.1 | 62% | 12% |
| Washington | 17-26wk ahead | -0.15 | -0.07 | 92% | 26% |
| Washington | peak intensity | -0.52 | -0.08 | 90% | 0% |
| Washington | peak week | -0.76 | -6.85 | 60% | 0% |
| Washington | total | -0.22 | -0.06 | 90% | 10% |
| Wyoming | all | -0.35 | -0.29 | 44% | 21% |
| Wyoming | 1-8wk ahead | -0.14 | -0.03 | 40% | 25% |
| Wyoming | 9-16wk ahead | -0.47 | -0.3 | 29% | 9% |
| Wyoming | 17-26wk ahead | -0.27 | -0.21 | 63% | 33% |
| Wyoming | peak intensity | -0.42 | -1.29 | 20% | 0% |
| Wyoming | peak week | -2.1 | -2.23 | 0% | 10% |
| Wyoming | total | -0.21 | -0.04 | 90% | 0% |

**Table S8.**
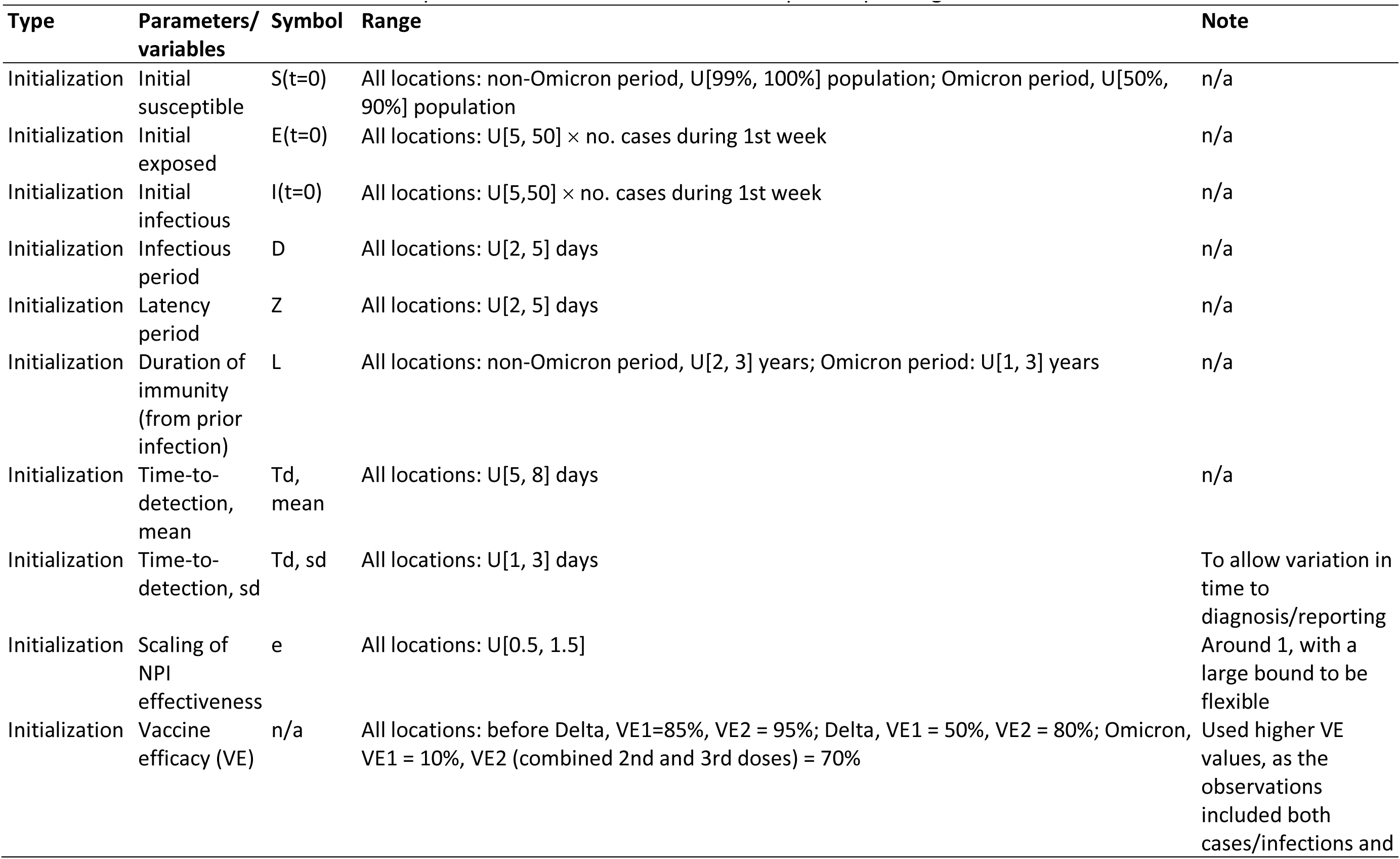
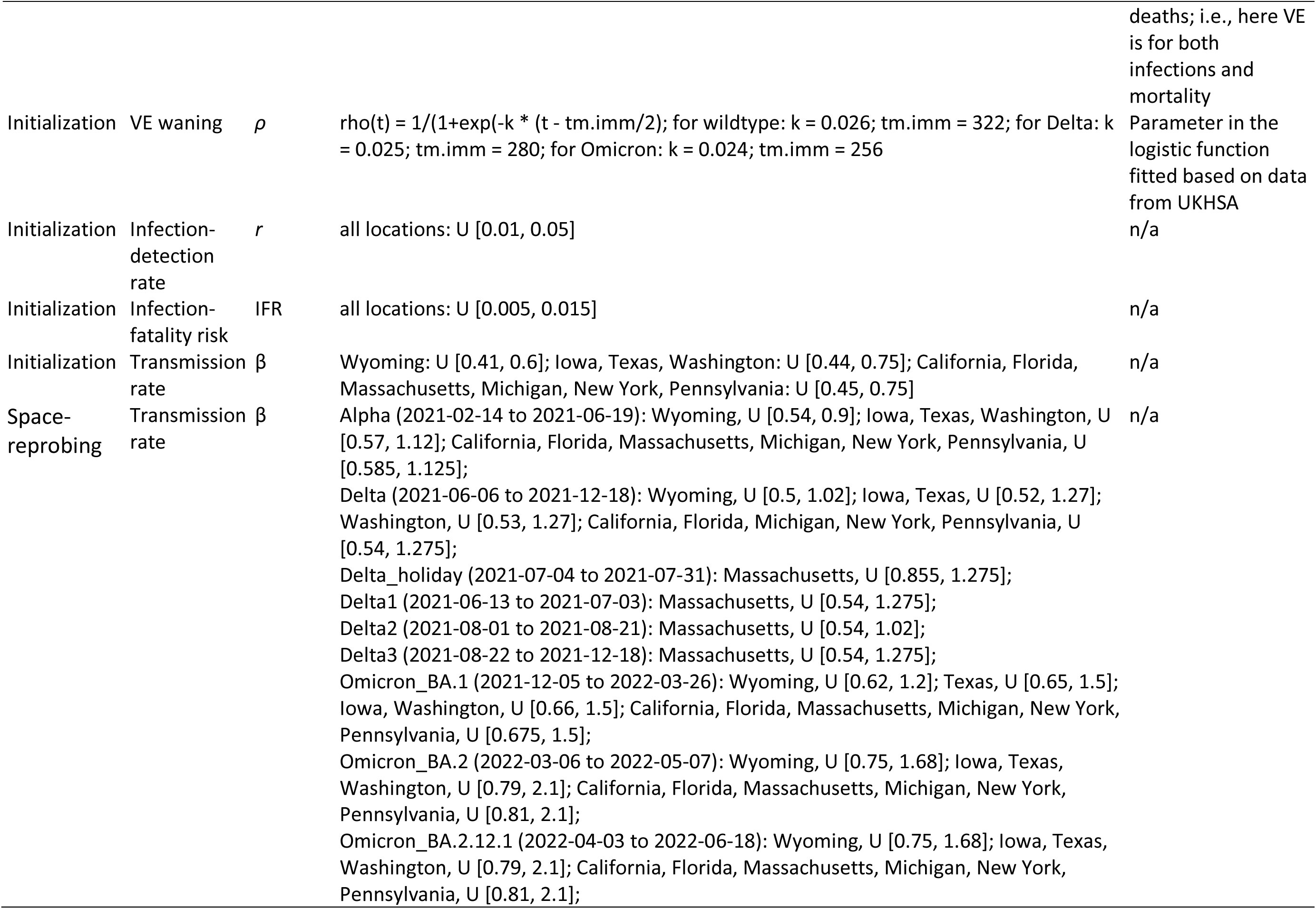
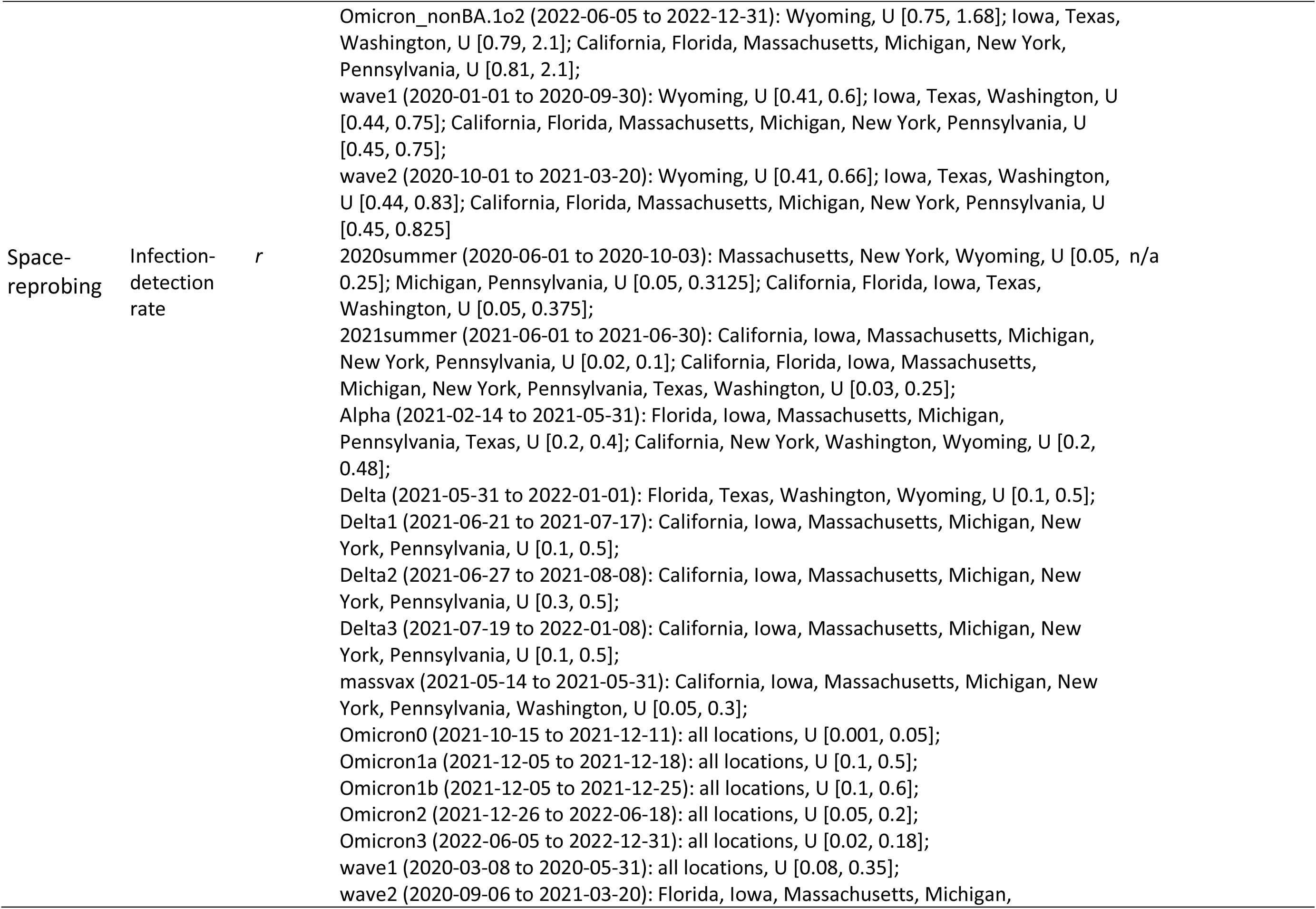
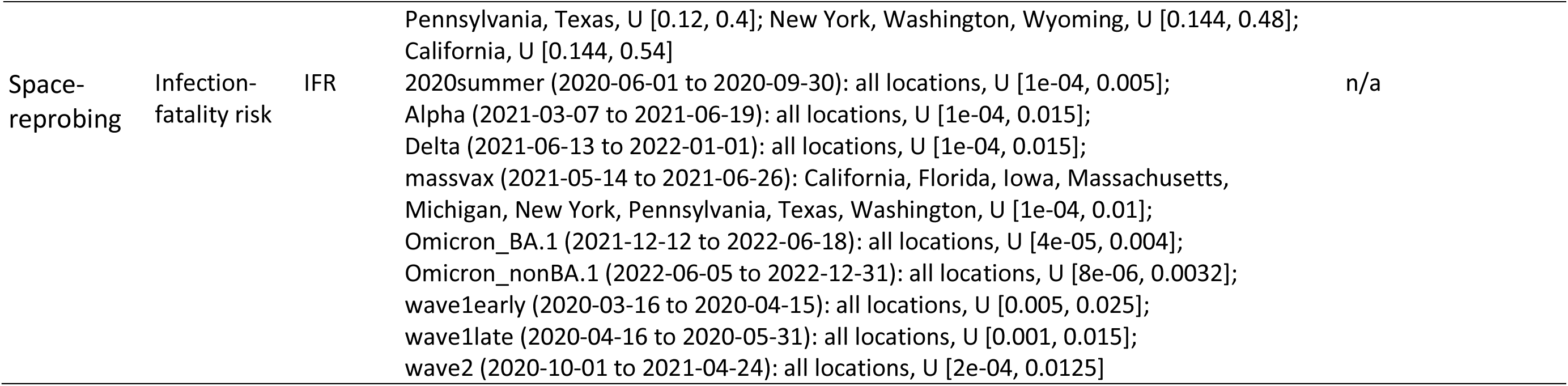
Prior ranges for the parameters and variables used in the model-inference system. Parameters/state variables are initialized by drawing from uniform distributions specified in the rows labeled “Initialization”. During the filtering process, space-reprobing is applied to explore the state space, i.e., a small fraction of the ensemble members are randomly replaced with values drawn from the uniform distributions specified in the rows labeled with “Space-reprobing”.

